# A type I IFN, prothrombotic hyperinflammatory neutrophil signature is distinct for COVID-19 ARDS

**DOI:** 10.1101/2020.09.15.20195305

**Authors:** Leila Reyes, Manuel A. Sanchez-Garcia, Tyler Morrison, Andrew J.M. Howden, Emily R. Watts, Simone Arienti, Pranvera Sadiku, Patricia Coelho, Ananda S. Mirchandani, Ailiang Zhang, David Hope, Sarah K. Clark, Jo Singleton, Shonna Johnston, Robert Grecian, Azin Poon, Sarah McNamara, Isla Harper, Max Head Fourman, Alejandro J. Brenes, Shalini Pathak, Amy Lloyd, Giovanny Rodriguez Blanco, Alex von Kriegsheim, Bart Ghesquiere, Wesley Vermaelen, Camila T. Cologna, Kevin Dhaliwal, Nik Hirani, David H. Dockrell, Moira K. B. Whyte, David Griffith, Doreen A. Cantrell, Sarah R. Walmsley

## Abstract

Acute respiratory distress syndrome (ARDS) is a severe critical condition with a high mortality that is currently in focus given that it is associated with mortality caused by coronavirus induced disease 2019 (COVID-19). Neutrophils play a key role in the lung injury characteristic of non-COVID-19 ARDS and there is also accumulating evidence of neutrophil mediated lung injury in patients who succumb to infection with SARS-CoV-2. We undertook a functional proteomic and metabolomic survey of circulating neutrophil populations, comparing patients with COVID-19 ARDS and non-COVID-19 ARDS to understand the molecular basis of neutrophil dysregulation. Expansion of the circulating neutrophil compartment and the presence of activated low and normal density mature and immature neutrophil populations occurs in ARDS, irrespective of cause. Release of neutrophil granule proteins, neutrophil activation of the clotting cascade and upregulation of the Mac-1 platelet binding complex with formation of neutrophil platelet aggregates is exaggerated in COVID-19 ARDS. Importantly, activation of components of the neutrophil type I interferon responses is seen in ARDS following infection with SARS-CoV-2, with associated rewiring of neutrophil metabolism, and the upregulation of antigen processing and presentation. Whilst dexamethasone treatment constricts the immature low density neutrophil population it does not impact upon prothrombotic hyperinflammatory neutrophil signatures.

**Summary:** Given the crucial role of neutrophils in ARDS and the evidence of a disordered myeloid response observed in COVID-19 patients, our work maps the molecular basis for neutrophil reprogramming in the distinct clinical entities of COVID-19 and non-COVID-19 ARDS.

**Conclusions:** This work maps the molecular basis for neutrophil reprogramming in the distinct clinical entities of COVID-19 and non-COVID-19 ARDS.

## Introduction

Coronavirus disease (COVID-19) is an acute respiratory condition caused by novel coronavirus (SARS-CoV-2) infection. In the most severe cases (termed “Critical COVID-19”), infection with SARS-CoV-2 can lead to the development of acute respiratory distress syndrome (ARDS) (1). ARDS is a clinical syndrome defined by the presence of bilateral pulmonary infiltrates on chest radiograph and arterial hypoxaemia that develops acutely in response to a known or suspected insult. ARDS is known to be the consequence of disordered inflammation (2), and is characterised by a protein-rich oedema in the alveoli and lung interstitium, driven by epithelial and vascular injury (2, 3) and increased vascular permeability (4, 5). Limited data exists regarding the mechanisms causing hypoxaemia and lung inflammation following infection with SARS-CoV-2, although post-mortem case reports provide evidence of diffuse alveolar damage, with the presence of proteinaceous exudates in the alveolar spaces, intra-alveolar fibrin and alveolar wall expansion (6). In previously described ARDS cohorts in which SARS-CoV-2 was not an aetiological factor, alveolar damage is associated with worsening hypoxia and increased mortality (7). In this context, hypoxia is a key driver of dysfunctional inflammation in the lung, augmenting neutrophil persistence and survival (8, 9) and promoting the release of pro-inflammatory mediators that cause ongoing tissue injury (2, 3). Non-dyspnoeic hypoxia is widely described in patients with severe COVID-19 (10), where it is associated with altered circulating leukocyte profiles with an increase in neutrophil to lymphocyte ratios and the presence of lymphopaenia (11, 12). More recently, post-mortem studies have revealed that the diffuse alveolar damage does not directly associate with the detection of virus, supporting the concept of aberrant host immune responses as drivers of tissue injury and pulmonary disease progression (13). A disordered myeloid response is further evidenced by analysis of gene clusters and surface protein expression of whole blood and peripheral blood mononuclear cell (PBMC) layers of patients with mild and severe COVID-19 (14). However, the functional relevance of these transcriptional signatures remains to be explored given the limited reliance of neutrophils on transcription to regulate protein expression (15). It also remains to be addressed whether the observed changes in neutrophil sub-populations are specific to COVID-19 ARDS or a reflection of the aberrant neutrophil inflammatory responses more broadly associated with the pathogenesis of ARDS. It is also unclear as to how these may be impacted by anti-inflammatory strategies including dexamethasone, which has shown to lower 28-day mortality for patients receiving invasive mechanical ventilation or oxygen alone (16).

One of the distinct features of COVID-19 that has emerged is the clinical evidence of a pro-thrombotic state, neutrophil retention within the lung microvasculature and the colocalisation of neutrophils with platelets in fibrin rich clots (17). Together with evidence of the formation of neutrophil extracellular traps (NETs) (18, 19) this raises the important question as to whether neutrophils are inappropriately activated within the circulation thus contributing to vascular injury and thrombosis in COVID-19. Exploring the differences in neutrophil responses between COVID-19 and non-COVID-19 ARDS provides an opportunity to understand the mechanisms common to ARDS and those that drive the hypercoagulable hyperinflammatory state specific to COVID-19 thus identifying urgently required therapeutic targets.

In this program of work, we compared the blood neutrophil populations of patients with COVID-19 ARDS to those of patients with non-COVID-19 ARDS, moderate COVID-19 and healthy controls to define the neutrophil host response to SARS-CoV-2. Prior to SARS-CoV-2, a significant confounder of ARDS studies has been the heterogeneity of the underlying processes that result in ARDS with hyperinflammatory and hypoinflammatory phenotypes described. Infection with SARS-CoV-2 provides a unifying trigger to this aberrant host response, whilst comparison between COVID-19 and non-COVID-19 ARDS allows us to identify neutrophil responses that are observed following infection with SARS-CoV-2, or associated with ARDS all cause.

## Methods

### Healthy donor and patient recruitment

Human peripheral venous blood was taken from healthy volunteers with written informed consent obtained from all participants prior to sample collection as approved by the University of Edinburgh Centre for Inflammation Research Blood Resource Management Committee (AMREC 15-HV-013). The collection of peripheral venous blood from patients diagnosed with COVID-19 and/or presenting with ARDS was approved by Scotland A Research Ethics Committee. Patient recruitment took place from April 2020 through January 2021 from The Royal Infirmary of Edinburgh, Scotland, UK through the ARDS Neut (20/SS/0002) and CASCADE (20/SS/0052) Study, with informed consent obtained by proxy. The presence of ARDS was defined using the Berlin criteria (20). Acute physiology and chronic health evaluation (APACHE II) score = acute physiology score + age points + chronic health points, was undertaken (minimum score = 0; maximum score = 71), where increasing score is associated with increasing risk of hospital death (21). Functional Comorbidity Index data (FCI) was also captured as an 18-item list of comorbidities used to adjust for the effect on physiological function (22). Scores were performed on intensive care unit admission or earliest time possible. Infection with SARS-CoV-2 was confirmed by polymerase chain reaction (PCR) either on nasopharyngeal swab, or deep airway samples. In non-COVID-19 ARDS viral infection was excluded.

### Isolation of human peripheral blood neutrophils

Up to 80 mL of whole blood was collected into citrate tubes. An aliquot of 5 mL of whole blood was treated with red cell lysis buffer (Invitrogen) and with the remaining volume, human blood leukocytes were isolated by dextran sedimentation and discontinuous Percoll gradients as described by Haslett et al. (1985) (23).

### Cell Culture

Normal density neutrophils (NDN) obtained from the polymorphonuclear (PMN) layer of healthy donors were resuspended at 5 × 10^6^/mL in Roswell Park Memorial Institute (RPMI) 1640 (Gibco) with 10% dialyzed foetal calf serum (Gibco) and 50 U/mL streptomycin and penicillin in normoxia (19 kPa) or hypoxia (3 kPa) at 5% CO_2_ as previously described (24). Cells were cultured in the absence or presence of interferon (IFN)α/ IFNβ (500 units/mL) and/or resiquimod (15 μM, Sigma-Aldrich) for the indicated time prior to harvest. For flow cytometry studies using dexamethasone and resiquimod, cells were cultured in hypoxia and pre-treated with varying doses of dexamethasone (0-1 μM) for 4 h, followed by treatment with resiquimod (15 μM) for 1 h. For heavy glutamine tracing studies, NDN were cultured in the presence of 2 mM U-13C5 glutamine (Cambridge). For extracellular flux analysis, cells were cultured in the presence or absence of interferon in hypoxia as above before transfer to an extracellular flux cell culture microplate after 3 hours.

### Flow cytometry

Lysed whole blood, PMN and PBMC layers isolated from Percoll gradients, as well as healthy control NDN used for dexamethasone/resiquimod studies were stained with Zombie Aqua™ Fixable viability dye (1:400) (Biolegend) to exclude dead cells from analysis. Cells were subsequently treated with Human TruStain FcX™ (1:100) (Biolegend) and stained for 30 min on ice with antibodies listed in Table 1 with appropriate fluorescence minus one (FMO) controls. Cells were then washed and fixed with 4 % paraformaldehyde (PFA) and acquired using BD LSRFortessa™ flow cytometer (Beckton Dickinson). Compensation was performed using BD FACSDiva™ software version 8.0 and data analysed in FlowJo version 10.2. Gating strategy to identify neutrophils, maturity and surface expression of various markers are outlined in Figure 1. Samples with neutrophil purity of <95% (CD66b+CD49d-) were excluded from analysis.

**Table 1.** List of antibodies used for multi-panel flow cytometry and microscopy. Detailed list of all antibodies used for flow cytometry and microscopy staining. Refer to the corresponding method details section for further information.

| **Antibody** | **Clone** | **Catalogue number** | **Fluorophore** | **Source** | **Concentration** |
| --- | --- | --- | --- | --- | --- |
| CD16 | eBioCB16 | 11-0168-42 | FITC | Ebioscience | 1:100 |
| CD63 | H5C6 | 353004 | PE | Biolegend | 1:100 |
| CD10 | K036C2 | 312214 | PE-Cy7 | Biolegend | 1:100 |
| CD66b | G10F5 | 305114 | AF700 | Biolegend | 1:100 |
| CD62L | DREG-56 | 304814 | APC-Cy7 | Biolegend | 1:100 |
| CD11b | M1/70 | 101243 | BV785 | Biolegend | 1:400 |
| CD49d | 9F10 | 304322 | BV421 | Biolegend | 1:100 |
| CD41 | HIP8 | 303729 | BV421 | Biolegend | 1:100 |
| CD18 | TS1/18 | 302117 | PE-Cy7 | Biolegend | 1:100 |

**Figure 1.**
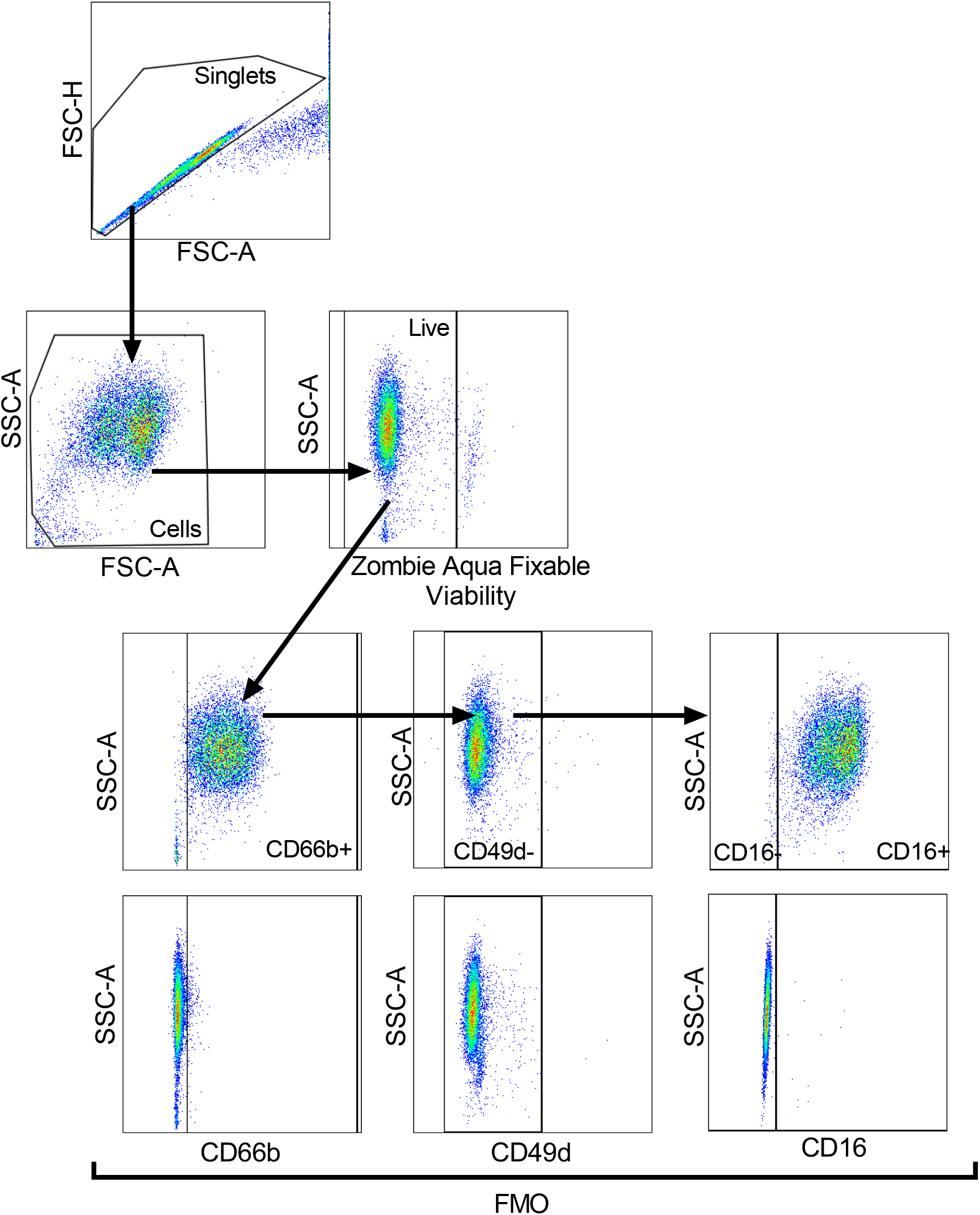
Representative plots of the gating strategy to analyse neutrophil populations. Strategy shown in the direction of the arrows. Cells were gated for singlets on a forward scatter height (FSC-H) vs. forward scatter area (FSC-A) plot. Singlets were then gated for cells on a side scatter area (SSC-A) vs. FSC-plot, with non-viable cells excluded according to SSC-A vs. Zombie Aqua Fixable Viability Dye parameter. Viable single cells were gated for CD66b+ cells to identify neutrophils and eosinophils excluded according to SSC-A vs. CD49d, with fluorescence minus one (FMO) controls used to set gates. CD66b+CD49d-cells (neutrophils) were gated for mature (CD16+) and immature (CD16-) neutrophils, with FMO controls used to set gate.

### Proteomic sample preparation

NDN were centrifuged at 300 × g for 5 min at 4 °C and resuspended in 7 mL of 0.2 % NaCl (w/v in H_2_O) for 5 min at room temperature and topped up with 7 mL of 0.2 % NaCl (w/v in H_2_O). Cells were washed twice in DPBS (Thermo Fisher), pelleted at 300 × g for 5 min at 4 °C and resuspended in 372 μL of freshly made 5% sodium dodecyl sulfate (SDS) lysis buffer and vortexed. Samples were then heat denatured in a heat block for 5 min at 100 °C and stored at –80 °C. Cell lysates were thawed and tris(2-carboxyethyl) phosphine hydrochloride (TCEP) and triethylammonium bicarbonate (TEAB) were added to a final concentration of 10 mM and 50 mM respectively. Lysates were shaken at 500 rpm at 22 °C for 5 min before being incubated at 98 °C for 5 min. Samples were allowed to cool and were then sonicated with a BioRuptor (30 cycles: 30 s on and 30 s off). Tubes were centrifuged at 17,000 × g to collect the cell lysate and 1 mL of benzonase (27.8 units) was added to each sample and samples incubated at 37 °C for 15 min. Samples were then alkylated with addition of 20 mM iodoacetamide for 1 h at 22 °C in the dark. Protein lysates were processed for mass spectrometry using s-trap spin columns following the manufacturer’s instructions (Protifi) (25). Lysates were digested with Trypsin at a ratio 1:20 (protein:enzyme) in 50 mM ammonium bicarbonate. Peptides were eluted from s-trap columns by sequentially adding 80 mL of 50 mM ammonium bicarbonate followed by 80 mL of 0.2 % formic acid with a final elution using 80 mL of 50 % acetonitrile + 0.2 % formic acid.

### LC-MS analysis

For each sample, 2 mg of peptide was analysed on a Q-Exactive-HF-X (Thermo Scientific) mass spectrometer coupled with a Dionex Ultimate 3000 RS (Thermo Scientific). LC buffers were the following: buffer A (0.1% formic acid in Milli-Q water (v/v)) and buffer B (80% acetonitrile and 0.1% formic acid in Milli-Q water (v/v)). 2 μg aliquot of each sample were loaded at 15 μL/min onto a trap column (100 μm × 2 cm, PepMap nanoViper C18 column, 5 μm, 100 Å, Thermo Scientific) equilibrated in 0.1% trifluoroacetic acid (TFA). The trap column was washed for 3 min at the same flow rate with 0.1% TFA then switched in-line with a Thermo Scientific, resolving C18 column (75 μm × 50 cm, PepMap RSLC C18 column, 2 μm, 100 Å). The peptides were eluted from the column at a constant flow rate of 300 nl/min with a linear gradient from 3% buffer B to 6% buffer B in 5 min, then from 6% buffer B to 35% buffer B in 115 min, and finally to 80% buffer B within 7 min. The column was then washed with 80% buffer B for 4 min and re-equilibrated in 3% buffer B for 15 min. Two blanks were run between each sample to reduce carry-over. The column was kept at a constant temperature of 50 °C at all times.

The data was acquired using an easy spray source operated in positive mode with spray voltage at 1.9 kV, the capillary temperature at 250 °C and the funnel RF at 60 °C. The MS was operated in DIA mode as reported earlier (26) with some modifications. A scan cycle comprised a full MS scan (m/z range from 350-1650, with a maximum ion injection time of 20 ms, a resolution of 120 000 and automatic gain control (AGC) value of 5 × 10^6^). MS survey scan was followed by MS/MS DIA scan events using the following parameters: default charge state of 3, resolution 30.000, maximum ion injection time 55 ms, AGC 3 x 10^6^, stepped normalized collision energy 25.5, 27 and 30, fixed first mass 200 m/z. The inclusion list (DIA windows) and windows widths are shown in Table 2. Data for both MS and MS/MS scans were acquired in profile mode. Mass accuracy was checked before the start of samples analysis.

**Table 2.**
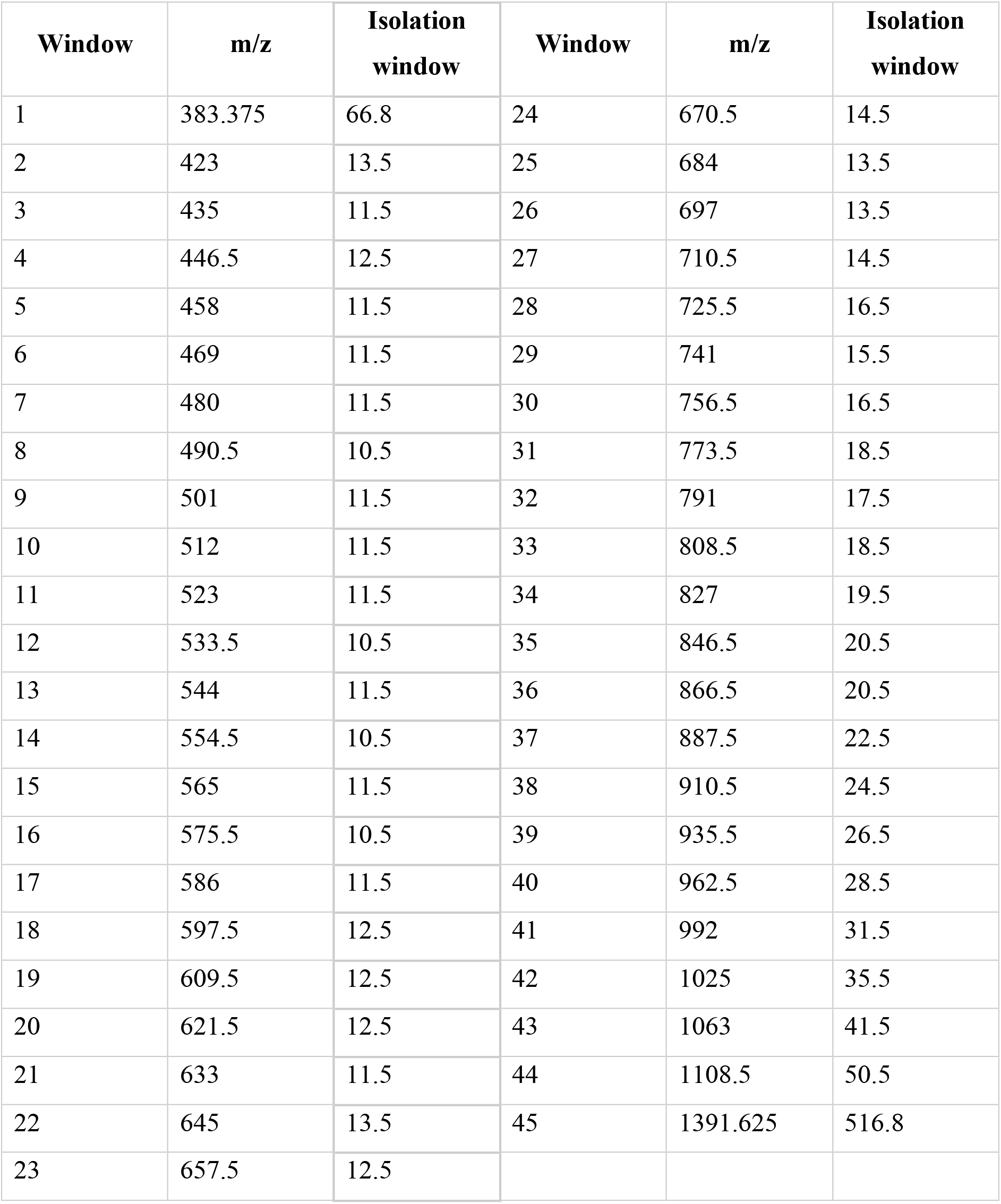
Inclusion list. Mass spectrometry isolation windows for data independent acquisition analysis. Refer to the corresponding method details section for further information.

### Analysis of proteomics data

The DIA data were analyzed with Spectronaut 14 using the directDIA option (27). Cleavage Rules were set to Trypsin/P, Peptide maximum length was set to 52 amino acids, Peptide minimum length was set to 7 amino acids and Missed Cleavages set to 2. Calibration Mode was set to Automatic. Search criteria included carbamidomethylation of cysteine as a fixed modification, as well as oxidation of methionine, deamidation of asparagine and glutamine and acetylation (protein N-terminus) as variable modifications. The FDR threshold was set to 1% Q-value at both the Precursor and Protein level. The single hit definition was to Stripped sequence. The directDIA data were searched against the human SwissProt database (July 2020) and included isoforms. The Major Group Quantity was set to the Sum of peptide quantity and the Minor Group Quantity was set to the Sum of the precursor quantity; Cross Run Normalization was disabled. Fold changes and P-values were calculated in R utilising the bioconductor package LIMMA version 3.7(28). The Q-values provided were generated in R using the “qvalue” package version 2.10.0. Estimates of protein copy numbers per cell were calculated using the histone ruler method (29). The mass of individual proteins was estimated using the following formula: CN × MW/NA = protein mass (g cell^−1^), where CN is the protein copy number, MW is the protein molecular weight (in Da) and NA is Avogadro’s Constant. Raw mass spectrometry data files and Spectronaut analysis files have been deposited to the ProteomeXchange (30) Consortium via the PRIDE (31) partner repository.

### Cell immunostaining for microscopy

PMN and PBMC layers isolated from Percoll gradients were fixed with 1.5 % PFA. FACS of NDN and LDN from PMN and PBMC layers respectively were performed using BD FACSAria™ Fusion flow cytometer fitted with a 70 µm nozzle and running BD FACSDiva™ software version 8.0 (Beckton Dickinson). Singlets were gated according to forward scatter height vs. forward scatter area (FSC-H vs. FSC-A) and side scatter height vs. side scatter area (SSC-H vs. SSC-A) parameters and NDN and LDN identified according to forward vs. side scatter (FSC vs. SSC) parameters. NDN and LDN were collected at 4 °C in 15 mL Falcon tubes pre-coated with Dulbecco’s phosphate-buffered saline (DPBS; Thermo Fisher).

Cells were pelleted and blocked with Fc Receptor Blocking Solution followed by staining with anti-CD41 antibody (Biolegend) and counterstaining with propidium iodide (Biolegend) according to manufacturer’s guidelines. Multichamber slides (Ibidi) were used to image the samples in a confocal microscope (Leica SP8). Image acquisition was performed at 63x magnification with the same settings across all images. Fiji software was used to process the images (32). Scale bars depict 5 μm.

### Measurement of granule protein levels

Enzyme-linked immunosorbent assay (ELISA) was performed according to manufacturer’s protocol to quantify MPO, lactoferrin and elastase levels (Abcam) in plasma from healthy donors, non-COVID-19 ARDS, COVID-19 patients and cell media supernatant of resiquimod treated NDN cultures from healthy donors (15 μM, Sigma-Aldrich).

### Metabolomic analysis

2.5 × 10^6^ neutrophils freshly isolated from the PMN layer of patients were centrifuged at 300 × g for 5 min at 4 °C, with pellets resuspended in 100 μL of 80% methanol. Alternatively, 5 × 10^6^ NDN from healthy controls were cultured in hypoxia for 4 h in the presence or absence of IFNα/IFNβ, washed twice in ice cold saline following culture and lysed in 200 μL of 50:30:20 methanol:acetonitrile:water. Following extraction, samples were stored at –80 °C. Relative metabolite abundance was determined using ion-pairing Reverse Phase High Performance Liquid Chromatography (RP-HPLC) or Hydrophilic Interaction Liquid Chromatography coupled to a Q-exactive Orbitrap Mass Spectrometer. Data were analysed in a targeted manner, using Xcalibur (Thermofisher Scientific) against an in-house compound library to integrate the area under the curve at the expected retention time. Individual metabolites were expressed relative to the mean of the healthy control population and analysed in Prism 9.00 (Graphpad Software Inc).

### Extracellular flux analysis

After culture, cells were harvested into sealed eppendorfs and maintained in hypoxia for one wash in warm saline. Cells were resuspended at 3 × 10^6^/mL in XF DMEM pH 7.4 (Agilent), supplemented with 2 mM glutamine and IFNα/IFNβ added to the appropriate cells at the concentrations described previously. 3 × 10^6^ neutrophils were adhered into each well of a 24-well cell culture microplate (Agilent) pretreated with cell-tak (Corning) to give triplicate samples per condition with 4 wells were left as media controls. After allowing CO_2_ to degas for 45 min in a hypoxic incubator, the plate was loaded into a Seahorse XFe 24 Analyzer (Agilent) operated in a hypoxic chamber (3% O_2_, 0.1% CO_2_; SCI-tive hypoxic work station, Baker Russkinn). Treatment compounds were resuspended in XF DMEM and cells were sequentially treated by injection of resiquimod (15*μ*M), glucose (10 mM, Sigma), oligomycin A (1 μM, Sigma) and 2-deoxyglucose (50 mM, Sigma). All media and compounds were pre-equilibrated in hypoxia. Data were acquired using Seahorse Wave Controller (version 2.6, Agilent) and analysed using Wave before exporting to GraphPad to pool for final analysis.

### Statistical analyses

Statistical tests were performed using Prism 9.00 software (GraphPad Software Inc). Data was tested for normality using Shapiro-Wilk test and outliers excluded according to Grubb’s test, with significance testing detailed in figure legends. Significance was defined as a p value of <0.05 after correction for multiple comparisons where applicable. Sample sizes are shown in figure legends, with each n number representing a different donor.

## Results

### Study population cohorts

To define the circulating neutrophil response to infection with SARS-CoV-2 we studied peripheral blood neutrophil populations isolated from hospitalised patients with moderate COVID-19 and COVID-19 ARDS, comparing these to critical care patients with non-COVID-19 ARDS and healthy controls (male, n = 4; female, n = 5; age range: 20 – 50 years) (Figure 2A). Patient demographic details are provided in Table 3. In accordance with the WHO COVID-19 classification, patients recruited had either moderate (clinical signs of pneumonia with oxygen saturations >90%) or critical (ARDS) COVID-19 (33). Patients with Berlin criteria ARDS had mean APACHE II scores of 18.1 (Non-COVID-19) and 14.8 (COVID-19) respectively. Viral infections were excluded from the aetiology of non-COVID-19 ARDS. Of the 12 patients recruited with COVID-19 ARDS, 9 received dexamethasone.

**Table 3.** Characteristics of study groups. For disease groups, all measurements were taken at the time of trial sample unless otherwise specified. Plus-minus values are means ± SD. ‡ For COVID-19 ARDS group, data provided for 9 patients that were receiving invasive mechanical ventilation (n=8) or non-invasive ventilation (n=1) at the time of the sample. The 3 other patients were receiving high flow nasal oxygen at the time of the sample, so these measurements were not available. Acute physiology and chronic health evaluation (APACHE II) score, ¶ Functional Comorbidity Index (FCI), arterial partial pressure of oxygen (PaO_2_), peripheral oxygen saturation (SpO_2_), positive end-expiratory pressure (PEEP).

| Characteristic | Non-COVID-19 ARDS<br>(n = 7) | COVID-19 ARDS<br>(n = 12) | Moderate COVID-19<br>(n = 3) |
| --- | --- | --- | --- |
| **Demographics** |  |  |  |
| Age – yr | 48.7±9.3 | 59.0±14.5 | 85.7±5.7 |
| Female sex – no. (%) | 5 (71.4) | 3 (25.0) | 2 (66.7) |
| Body-mass index – kg/m<sup>2</sup> | 27.7±2.9 | 33.1±8.5 | 19.9±3.3 |
| **Clinical Data** |  |  |  |
| Cause of ARDS |  |  |  |
| Pulmonary – no. (%) | 5 (71.4) | 12 (100.0) |  |
| Extra-pulmonary | 2 (28.6) | 0 (0.0) |  |
| APACHE II Score\|\| | 18.1±5.4 | 14.8±3.9 |  |
| Functional Comorbidity Index¶ | 1.4±0.8 | 1±0.7 |  |
| Lowest SpO<sub>2</sub> in 6 hrs prior to ICU admission | 89.4±6.0 | 77±21 |  |
| Organ Injury and Support |  |  |  |
| Invasive mechanical ventilation – no. (%) | 7 (100.0) | 8 (66.7) |  |
| Renal replacement therapy – no. (%) | 1 | 0 |  |
| Vasopressors – no. (%) | 7 (100.0) | 4 (33.3) |  |
| **ARDS Therapy‡** |  |  |  |
| Peak pressure (cmH<sub>2</sub>O) | 25.0±7.2 | 24±6 |  |
| PEEP (cmH<sub>2</sub>O) | 7.9±3.2 | 10±4 |  |
| Tidal volume (ml) | 366.0±59.5 | 402±63 |  |
| Respiratory rate (breaths/min) | 28.12±2.3 | 23.1±3.8 |  |
| **Medications** |  |  |  |
| Lopinavir-Ritonavir – no. (%) | 0 (0.0) | 3 (25.0) | 0 (0.0) |
| Hydroxychloroquine – no. (%) | 0 (0.0) | 2 (16.7) | 0 (0.0) |
| Dexamethasone – no. (%) | 0 (0.0) | 9 (75.0) | 0 (0.0) |

**Figure 2.**
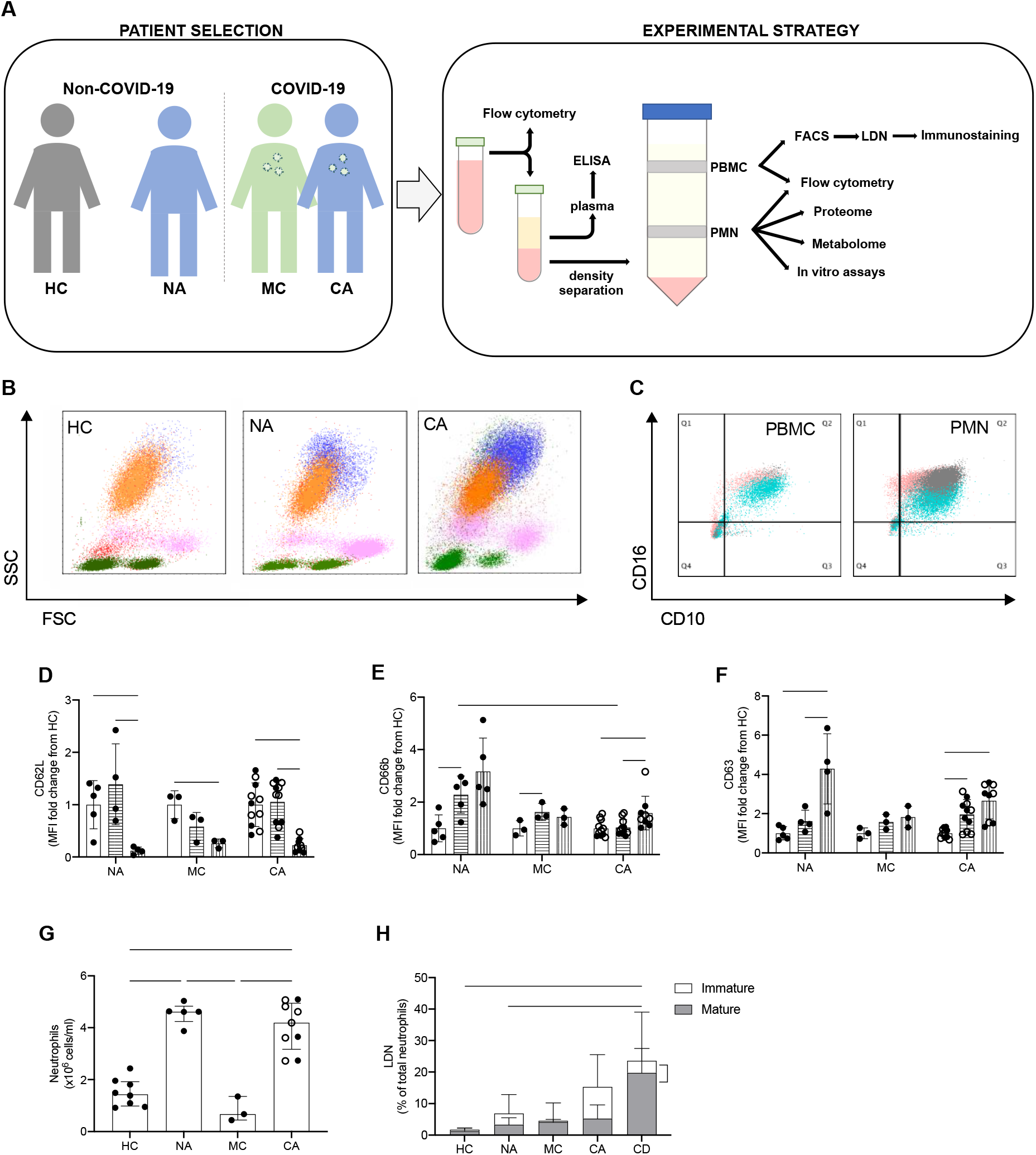
Circulating neutrophil populations are expanded in COVID-19 and non-COVID-19 ARDS. (A) Patient classification (healthy control, HC, non-COVID-19 ARDS, NA, moderate COVID-19, MC, and COVID-19 ARDS, CA), neutrophil isolation, and workflow depicted. (B) Representative side scatter (SSC) vs. forward scatter (FSC) plots of stained whole blood from HC, NA and CA displaying lymphocyte (green), monocyte (pink), mature (CD16+, orange) and immature (CD16-, blue) neutrophil populations. (C) Representative CD16 vs. CD10 dot plots of stained polymorphonuclear (PMN) and peripheral blood mononuclear cell (PBMC) layers isolated by Percoll gradients from HC (grey), NA (blue) or CA (pink) patients, with quadrant2 (Q2) delimiting the CD16+CD10+ (mature neutrophils) area. (D-F) Surface expression of neutrophil activation markers expressed as a fold change of geometric mean fluorescence intensity (MFI) from normal density neutrophils (NDN) respective to the disease state as determined by flow cytometry analysis of mature NDN (CD66b+CD16+, open bars), mature low density neutrophils (LDN) (CD66b+CD16+, horizontal striped bars) and immature LDN (CD66b+CD16-, vertical striped bars) from NA (n = 5), MC (n = 3), or COVID-19 (n = 11; open circles, dexamethasone treated patients) patients. Data are mean ± SD. ∗p < 0.05, determined by repeated two-way ANOVA and Sidak’s post hoc-testing. (G) Total neutrophil counts of HC (n = 8), NA (n = 5), MC (n=3) and CA (n = 11) performed by haemocytometer and differential cell count established by flow cytometry. Data are mean ± SD. ∗∗∗∗p < 0.0001, determined by one-way ANOVA and Holm-Sidak’s post hoc-testing. (H) The proportion of mature (CD66b+CD16+CD10+, grey bars) and immature (CD66b+CD16-CD10-, white bars) LDN isolated from patient cohorts as described in (G), with CA patients treated with dexamethasone as CD, were measured by flow cytometry. Data are mean ± SD. ∗p < 0.05, ∗∗∗p < 0.001, determined by repeated measures two-way ANOVA and Sidak’s post hoc-testing.

### Circulating neutrophil populations are expanded in COVID-19 and non-COVID-19 ARDS

To explore the different neutrophil populations, flow cytometry analysis of whole blood was first performed to identify CD66b+ cells as neutrophils, with CD16 used as a marker of maturity. CD66b+CD16+ and CD66b+CD16-cells were observed, indicating the presence of a heterogenous population of mature and immature neutrophils in ARDS patients, regardless of COVID-19 status (Figure 2B). Given immature neutrophils are characteristically low-density neutrophils (LDN) and associated with disease (34), flow cytometry analysis was performed on PMN and PBMC layers isolated using Percoll density gradients. Further characterisation of neutrophil maturity was undertaken by CD10 expression and showed both a mature (CD66b+CD16+CD10+) and immature (CD66b+CD16-CD10-) LDN population in the PBMC layer of non-COVID-19 and COVID-19 ARDS patients (Figure 2C). In contrast, these populations are notably absent in the PBMC layer of healthy control individuals (Figure 2C). Importantly, these LDN populations demonstrated evidence of increased activation states with loss of CD62L (Figure 2D), and upregulation of both CD66b and CD63 (Figure 2E-F). Total neutrophil counts generated from Percoll preparations showed a large expansion of neutrophils in ARDS (Figure 2G). Though a major proportion of the neutrophil population consisted of mature NDN from the PMN layer, we detected the presence of immature and mature low density neutrophil populations in ARDS patients (Figure 2H). Of note, the increase in immature LDN in the COVID-19 ARDS cohort was significantly reduced in those receiving dexamethasone despite a retained expansion of NDN and mature LDN populations (Figure 2H).

### Circulating neutrophils restructure their proteomes whilst retaining global cellular processes in COVID-19 and non-COVID-19 ARDS

A growing body of studies have described a disordered myeloid response following infection with SARS-CoV-2 using single cell RNA sequencing (scRNA-seq). These studies provide important insights to the reprogramming of myelopoiesis and the emergence of precursor neutrophil populations. However, there is a real need to understand how these transcriptional signatures relate to functional changes in myeloid cell responses, which requires information at a protein level. To understand changes in the functional proteome of circulating neutrophils we used a label free Data Independent Acquisition (DIA) mass spectrometry approach. Estimates of protein copy numbers per cell were calculated using the histone ruler method (29), along with total cellular protein content and the mass of subcellular compartments. We compared protein abundance between non-COVID-19 ARDS, COVID-19 ARDS and healthy control neutrophil populations. Analysis of the NDN populations, common to both healthy control and ARDS identified around 4500 proteins (Figure 3A), with a subtle reduction in the total protein content of COVID-19 ARDS neutrophils (Figure 3B). We observed preservation of global cellular processes across all disease groups evidenced by equivalent mitochondrial protein content, ribosomal protein content, nuclear envelope protein abundance and cytoskeletal protein abundance (Figure 3C). Key components of the translation initiation complex were also conserved across health and disease groups (Figure 3D). This would suggest that any differences observed in key neutrophil functions are not driven by a loss of core cellular processes and, therefore, more likely to be consequent upon activation of signalling pathways in response to infectious and inflammatory challenges. Globally we only detected a small number of proteins involved in transcription factor activity whose abundance was modified in response to COVID-19 (Figure 3E). These included the interferon regulated proteins TRIM22 and STAT1, which were induced in COVID-19 ARDS neutrophils and the glucocorticoid receptor NR3C1 which was down regulated.

**Figure 3.**
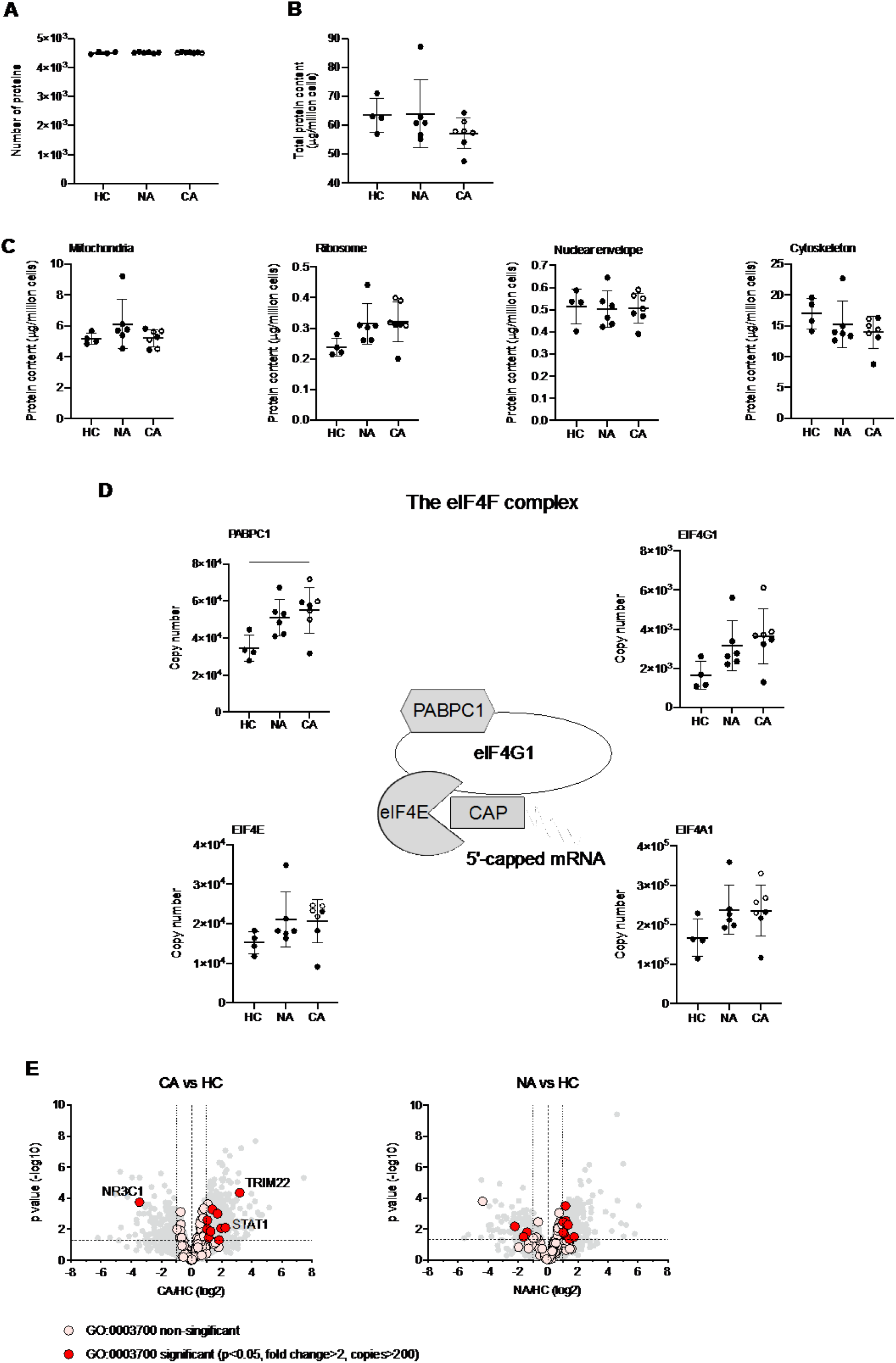
Circulating neutrophils preserve global cellular processes in both COVID-19 and non-COVID-19 ARDS. (A) Number of proteins identified by proteomic analysis in normal density neutrophils (NDN) isolated from healthy controls (HC, n = 4), non-COVID-19 ARDS patients (NA, n = 6) and COVID-19 ARDS patients (CA, n = 7), with open circles depicting the data corresponding to dexamethasone-treated patients. Refer to methods section for details. Data are mean ± SD. (B-C) Total protein content, protein content of mitochondria (GO:0005739), ribosomes (Kyoto Encyclopaedia of Genes and Genomes annotation 03010), nuclear envelope (GO:0005635) and cytoskeleton (GO:0005856) in the same samples described in (A) determined by proteomic analysis. Data as mean ± SD. (D) Abundance of components of the eIF4F translation initiation complex (figure adapted from Howden et al. 2019(49)) in the same samples described in (A) determined by proteomic analysis. Data as mean ± SD. (E) Volcano plots reflecting the expression profile of transcription factors in the samples from CA vs. HC, and NA vs. ARDS depicted in (A) after proteomic analysis. Proteins were included with the annotation GO:0003700 (DNA binding transcription factor activity). Horizontal dashed line indicates a P value = 0.05, outer vertical dashed lines indicate a fold change = 2. P values calculated using Linear Models for Microarray data analysis. The DNA binding transcription factor proteins TRIM22, STAT1 and NR3C1 are highlighted.

To determine which components of the neutrophil proteome remodel in patients with COVID-19 and non-COVID-19 ARDS we undertook Linear Models for Microarray data (LIMMA) analysis to identify significant differences in protein abundance (data are available via ProteomeXchange). We identified more than 200 proteins to be increased in abundance between COVID-19 ARDS and healthy control neutrophils which were not significantly changing in non-COVID ARDS (Figure 4A-B). Gene ontology (GO) term enrichment analysis of these differentially regulated proteins identified a COVID-19 signature which was defined by a greater abundance of proteins in type I IFN signalling pathways and the platelet degranulation (Figure 4B). Change in expression of cullin 2, cyclin dependent kinase 2, minichromosome maintenance complex components (MCM3-5 and MCM7) and phosphoribosyl pyrophosphate amidotransferase, proteins associated in other cell types with cell cycle control, was common to both COVID-19 and non-COVID-19 ARDS, whilst proteins important for mitochondrial translational termination and cell surface receptor signalling pathways were enriched in non-COVID-19 ARDS samples (Figure 4B). We identified 115 proteins with reduced abundance in ARDS (all cause) versus healthy control neutrophils, including some proteins that were specific to COVID-19. However, distinct biological processes impacted by SARS-CoV-2 infection were not identified among those proteins with reduced abundance.

**Figure 4.**
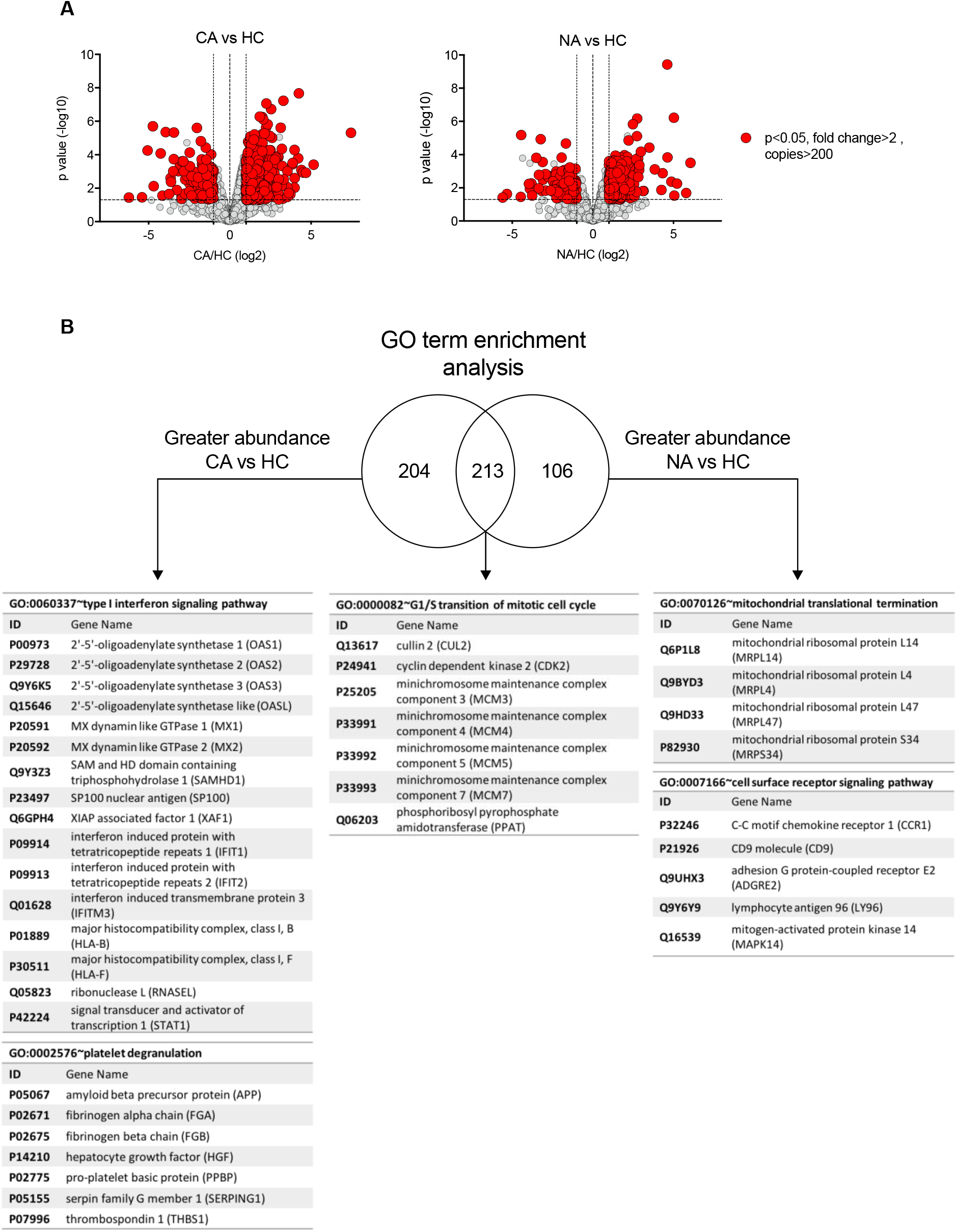
Specific proteome remodelling in circulating neutrophils in response to COVID-19 ARDS and ARDS. (A) Volcano plots obtained from proteomic survey of normal density neutrophils (NDN) isolated from healthy controls (HC, n = 4), non-COVID-19 ARDS patients (NA, n = 6) and COVID-19 ARDS patients (CA, n = 7). Refer to methods section for details. Proteins with a P value <0.05 (horizontal dashed lines), fold change >2 (outer vertical dashed lines) and a copy number >200 in at least one condition after Linear Models for Microarray data analysis were considered as significantly different in the comparisons CA vs HC (left) and NA vs HC (right). (B) GO term enrichment analysis for proteins significantly increased in abundance in CA and NA patients vs HC. Venn diagram shows the numbers of proteins uniquely increased in abundance in CA and NA and also the number of proteins shared between these 2 groups. A selection of the top enriched GO terms and the corresponding proteins are shown.

### COVID-19 ARDS neutrophils form aggregates with platelets and activate prothrombotic pathways with enrichment in the low density population

A striking clinical and post-mortem observation in patients with COVID-19 is the prevalence of micro and macrovascular thrombosis. With previous evidence of NET formation (18, 19), together with the colocalization of neutrophils with platelets in fibrin rich clots and our identification of a platelet degranulation signature within the COVID-19 ARDS samples, this led us to question the mechanism by which neutrophils could be contributing to immune mediated thrombosis in COVID-19. NDN displayed an overall increase in proteins associated with fibrin clot formation; fibrinogen alpha, fibrinogen beta and coagulation factor XIII alpha chain (F13A1) and a failure to induce proteins that inhibit fibrin clot formation in NDN like antithrombin-III (Figure 5A). This signature was greatest in COVID-19 ARDS neutrophils (Figure 5A). We also detected a platelet protein signature indicated by the presence of the platelet proteins platelet factor 4, platelet basic protein (Figure 5B). Confocal imaging on sorted mature neutrophil populations from COVID-19 ARDS patients subsequently revealed the existence of a direct physical association between LDN and platelets in these patients, as opposed to neutrophils from healthy donors (Figure 5C). To understand how neutrophil platelet aggregates were forming we looked for evidence of platelet activation on the neutrophil surface, and neutrophil expression of adhesion molecules involved in platelet interactions. Initial measurements for expression of CD41, a marker of platelet activation, revealed the presence of CD41 on mature LDN isolated from COVID-19 patients (Figure 5D). This coincided with a significant increase in mature neutrophil expression of the CD11b component of the Mac-1 platelet binding complex (Figure 5E). This phenotype was specific to the mature neutrophil populations, with only low-level surface expression of CD41, CD18, CD11b observed in the immature LDN population (Figure 5D-E).

**Figure 5.**
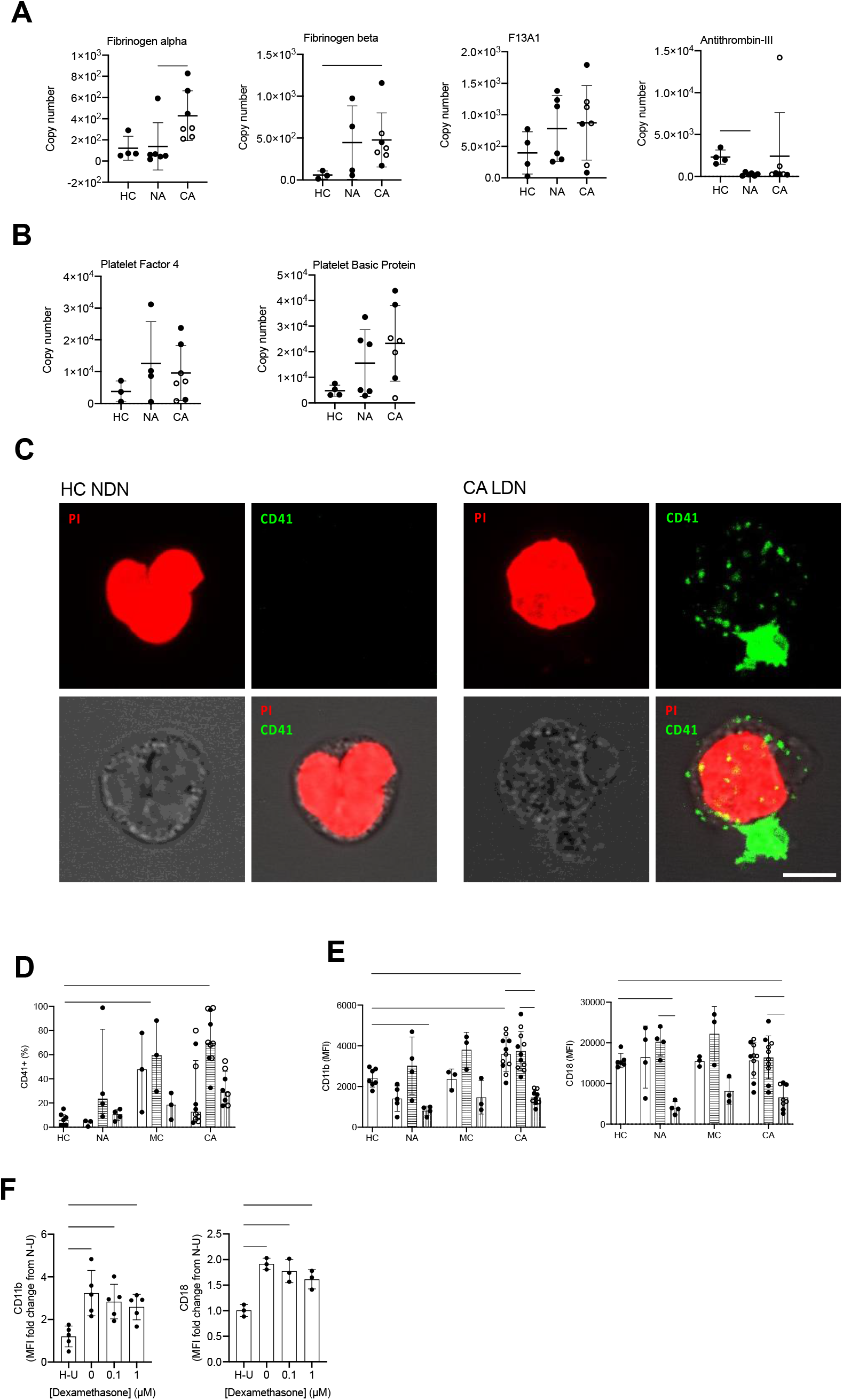
COVID-19 ARDS neutrophils form aggregates with platelets and activate prothrombotic pathways. (A) Copy numbers of proteins that regulated fibrin clot formation in normal density neutrophils (NDN) isolated from healthy controls (HC, n = 3-4), non-COVID-19 ARDS (NA, n = 4-6), and COVID-19 ARDS (CA, n = 7, with open circles depicting the data corresponding to dexamethasone-treated patients) patients determined by proteomic analysis. For fibrinogen alpha, fibrinogen beta and antithrombin-III, data as mean ± SD, ∗p < 0.05, determined by Kruskal-Wallis and Dunn’s post hoc-testing. For coagulation factor XIII alpha chain (F13A), data as mean ± SD. (B) Copy numbers of proteins associated with platelets in the same samples described in (A) determined by proteomic analysis. Data as mean ± SD. (C) Representative confocal images from NDN obtained from a healthy donor and LDN from a CA patient isolated by FACS and stained for propidium iodide (top left panel, red) and CD41 (top right panel, green). Bright field image was used to delimit cell contour (bottom left panel, grey scale). A composite image is shown in bottom right panel. Scale bar corresponds to 5 μm, 63x magnification. (D) Percentage of NDN (open bars), mature LDN (horizontal striped bars) and immature LDN (vertical striped bars) isolated from HC (n = 6-7), NA (n = 3-5), MC (n = 3) or CA patients (n = 8-11; open circles depict dexamethasone treated patients) with surface expression of CD41. Data are median ± I.Q.R. ∗p < 0.05, ∗∗∗∗p < 0.0001 vs. HC, determined be Kruskal-Wallis and Dunn’s post hoc-testing. (E) Surface expression of CD11b and CD18 (Mac-1 complex) displayed as geometric mean fluorescence intensity (MFI) determined by flow cytometry analysis of neutrophil populations described in (D). Data are mean ± SD. ∗∗p < 0.01, ∗∗∗p < 0.001, determined by repeated measures two-way ANOVA and Sidak’s post hoc-testing; ∗p < 0.05, ∗∗p < 0.01 vs. HC, determined be one-way ANOVA and Holm-Sidak’s post hoc-testing. (F) Surface expression of CD11b and CD18 (Mac-1 complex) expressed as a fold change of MFI of HC NDN cultured under untreated normoxic conditions (N-U) determined by flow cytometry analysis of HC NDN cultures in hypoxia under untreated conditions (H-U) or with varying doses of dexamethasone for 3 h and follow-up treatment with resiquimod for 1 h. Data are mean ± SD (n = 3-5). ∗p < 0.05, determined by repeated measures one-way ANOVA and Holm-Sidak’s post hoc-testing.

Toll like receptors (TLRs) are important for viral recognition by the innate immune response. TLR family members 7 and 8 have been previously reported to enable recognition of single stranded RNA viruses including influenza and SARS-CoV-2 (35, 36). To directly address whether neutrophil sensing of SARS-CoV-2 RNA was important for the regulation of Mac-1, we stimulated healthy control neutrophils with TLR 7 and 8 agonist resiquimod (37). Additionally, hypoxic culture conditions were used to replicate the systemic hypoxia that circulating neutrophils are exposed to in patients with COVID-19 ARDS. Resiquimod up-regulated neutrophil expression of both components of the Mac-1 platelet binding complex, CD11b and CD18 (Figure 5F) replicating the observed phenotype of COVID-19. In keeping with the patient data, the addition of dexamethasone to resiquimod stimulation did not impact CD11b or CD18 expression (Figure 5F).

The presence of neutrophil platelet aggregates in patients with COVID-19 ARDS led us to question why neutrophils were binding to activated platelets, and whether there was evidence that neutrophils themselves were becoming inappropriately activated in the blood. Neutrophils express a plethora of cell surface receptors to enable them to respond to noxious stimuli. A key element of this response is the highly regulated release of cytotoxic granule proteins. However, inappropriate degranulation in the lung tissue during ARDS is associated with epithelial and vascular damage which in turn potentiates lung injury (38). In health, the release of toxic granules by neutrophils in the circulation is limited by the requirement of a second activation stimulus following neutrophil priming (39). Comparison of the proteomes of NDN populations revealed that granule cargo proteins are highly abundant and account for approximately 20% of the neutrophil protein mass (Figure 6A). In both COVID-19 and non-COVID-19 ARDS, whilst we observed an equivalent abundance of primary (CD63, CD68 and Presenilin-1), secondary (Ras related proteins 1A-B and 2A-C), secondary and tertiary (secretory carrier membrane protein 1-4, vesicle associated membrane protein 2) and specifically tertiary (solute carrier 11A1) granule membrane proteins (data are available via ProteomeXchange), there is a relative reduction in the abundance of the granule cargo proteins within these circulating cells (Figure 6B). Survey of these individual proteases reveals these changes to be modest, but to occur across the different granule compartments and to be amplified in COVID-19 (Figure 6C-J). To address whether this relative reduction in intra-cellular granule protein content was consequent upon neutrophil degranulation, we quantified surface expression of CD63, a protein known to be externalised upon degranulation, and CD66b, whose surface expression augments in response to degranulation. We observed a significant increase in CD63 and CD66b expression which was specific to the COVID-19 ARDS neutrophils (Figure 6K). Importantly an increase in serum levels of the neutrophil granule proteins myeloperoxidase (MPO), lactoferrin and elastase in the COVID-19 ARDS patient cohort (Figure 6L) confirmed a phenotype of enhanced circulating neutrophil degranulation in the COVID-19 ARDS patient cohort, which was not impacted by dexamethasone (Figure 6M). Consistent with neutrophil sensing of SARS-CoV-2 RNA promoting activation and degranulation, stimulation of healthy control neutrophils with the TLR7 and 8 agonist resiquimod increased neutrophil shedding of CD62L and upregulated expression of CD66b and CD63 (Figure 6N). This resulted in an increase in detectable levels of the granule proteins MPO and lactoferrin in the cell culture supernatants (Figure 6O).

**Figure 6.**
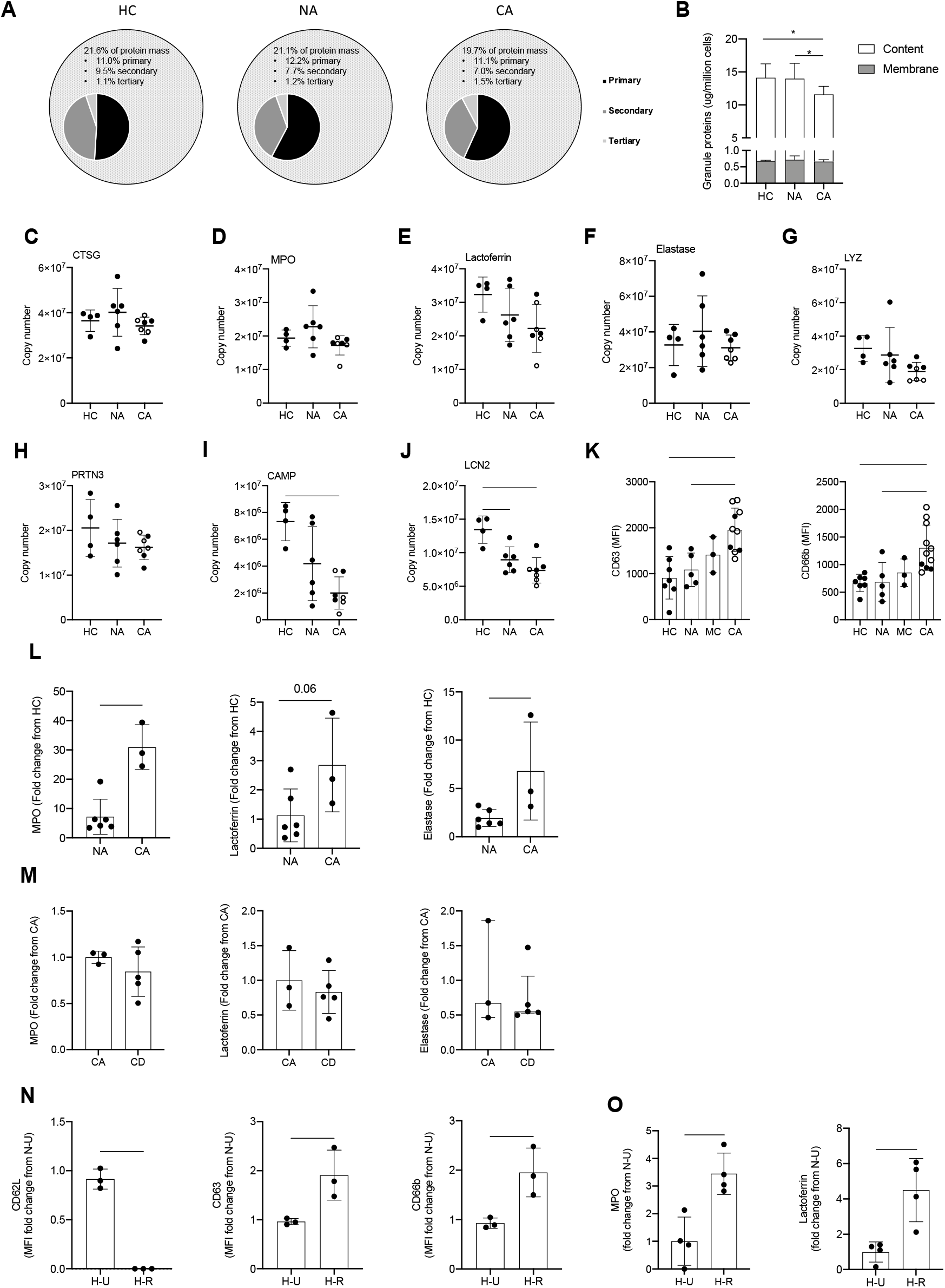
Enhanced circulating neutrophil degranulation in COVID-19. (A) Pie charts show distribution of protein mass for proteins integrating primary, secondary or tertiary granules in healthy control (HC, n = 4), non-COVID-19 ARDS (NA, n = 6) and COVID-19 ARDS (CA, n = 7) patients. Data obtained from proteomic analysis of normal density neutrophils (NDN). (B) Membrane (grey bars) and content (white bars) granule cargo protein abundance in NDN isolated from the samples described in (A). Data are mean ± SD. ∗p < 0.05, determined by two-way ANOVA and Tukey’s post hoc-testing. (C-J) Copy numbers of granule proteins in the samples described in (A, open circles depict dexamethasone-treated patients). For (C-E), data are mean ± SD. For (F-J), data are mean ± SD. ∗∗p < 0.01, determined be one-way ANOVA and Holm-Sidak’s post hoc-testing. (K) Surface expression of CD63 and CD66b displayed as mean fluorescence intensity (MFI) determined by flow cytometry analysis of NDN isolated from HC (n = 7), NA (n = 5), moderate COVID-19 (MC, n = 3) and CA (n = 12, open circles depict dexamethasone-treated patients) patients. For CD63, data are mean ± SD. ∗∗p < 0.01, ∗∗∗p < 0.001, determined be one-way ANOVA and Holm-Sidak’s post hoc-testing. For CD66b, data are median ± I.Q.R. ∗p < 0.05, ∗∗p < 0.01, determined by Kruskal-Wallis and Dunn’s post hoc-testing. (L) Granule protein levels in serum of NA (n = 6), and CA (n = 3) patients measured by ELISA represented as a fold change from HC. For MPO, data are median ± I.Q.R. ∗p < 0.05, determined by Mann-Whitney test. For elastase, data are mean ± SD. ∗p < 0.05, determined by unpaired t-test. (M) Granule protein levels in serum of CA (n = 3), and COVID-19 ARDS dexamethasone-treated (CD, n = 5) patients measured by ELISA represented as a fold change from CA. For MPO and lactoferrin, data are mean ± SD. For elastase, data are median ± I.Q.R. (N) Surface expression of activation markers expressed as a fold change of MFI of HC NDN under untreated normoxic conditions (N-U) determined by flow cytometry analysis of HC NDN cultures in hypoxia under untreated conditions (H-U) or with resiquimod (H-R) for 1 h. Data are mean ± SD. ∗p < 0.05, ∗∗p < 0.01, determined by paired one-tailed t-test. (O) Granule protein levels measured in H-U and H-R HC NDN culture supernatants at 4 h by ELISA expressed as a fold change of a N-U group. Data are mean ± SD. ∗p < 0.05, determined by paired t-test.

### Activation of neutrophil type I interferon signalling pathways and antigen presentation in COVID-19

Type I IFN are a group of cytokines which characterise the anti-viral response but are also implicated in inflammatory disease and in malignancy. Persistent high levels of circulating type I IFN are associated with more severe disease in the late stages of disease (40), thought to be due to dysfunctional inflammation rather than uncontrolled viral infection. This complexity is further reflected in the divergent signals in IFN stimulated genes (ISG) described in whole blood and PBMC myeloid cell populations, where select changes in transcript abundance are used to identify specific cell clusters (14). Here, we report using LIMMA analysis of NDN proteomes a type I IFN signature within the COVID-19 ARDS neutrophils (Figure 4B). We therefore surveyed the abundance of proteins involved in anti-viral responses downstream of IFNα/β receptor (IFNAR). This revealed across the pathway a greater abundance of proteins important for type I IFN signalling and anti-viral responses in COVID-19 ARDS neutrophils including 2’,5’-oligoadenylate synthetase (OAS) proteins which activate RNase L (Figure 7A), Eukaryotic Translation Initiation Factor 2-alpha Kinase 2 (EIF2AK) which inhibits viral transcription (Figure 7B) and the GTP binding Mx proteins which inhibit viral replication (Figure 7C).

**Figure 7.**
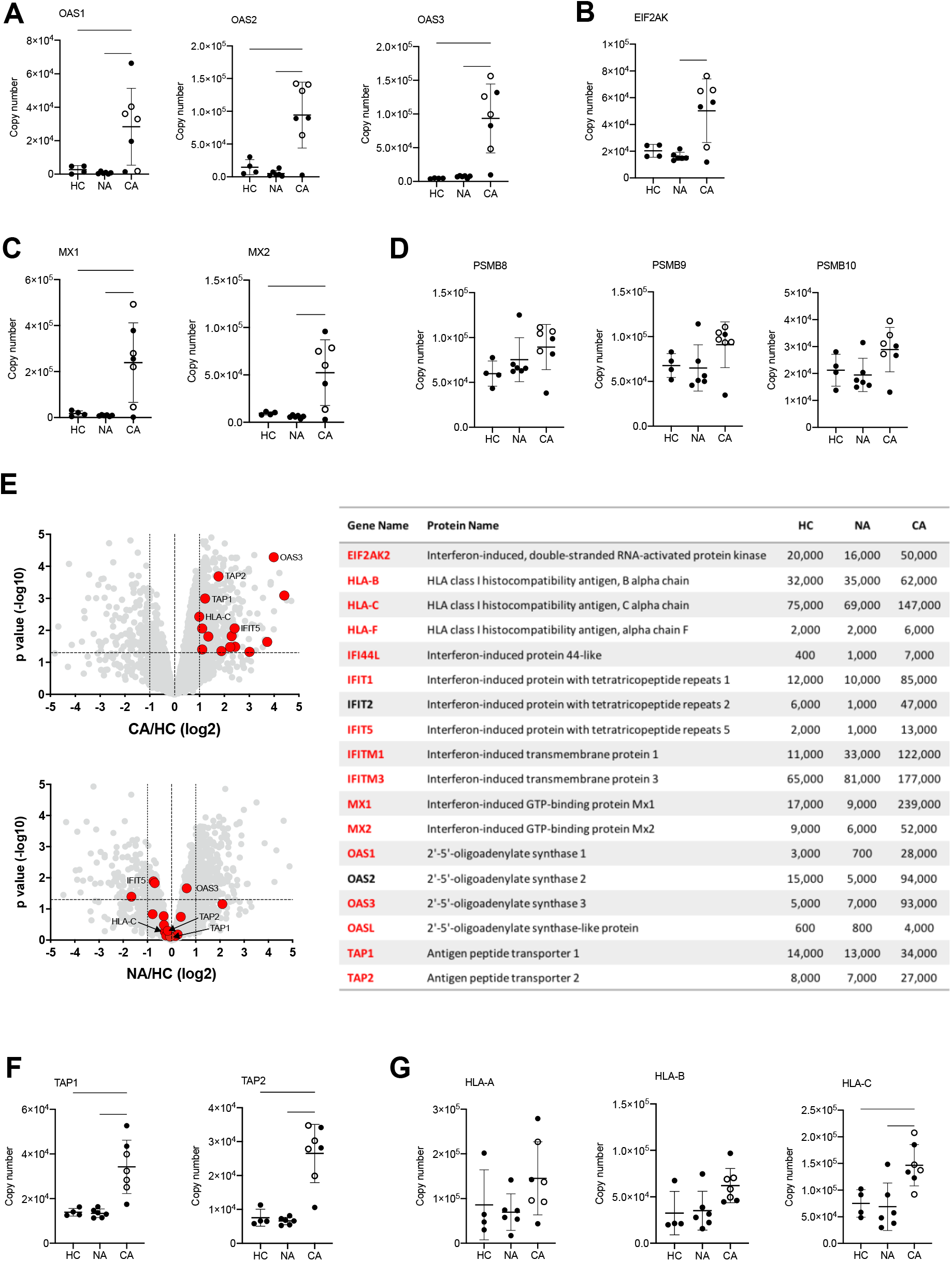
Activation of neutrophil type I interferon signalling pathways and antigen presentation in COVID-19. (A) Copy numbers of OAS proteins involved in type I IFN signalling and anti-viral responses in normal density neutrophils (NDN) isolated from healthy controls (HC, n = 4), non-COVID-19 ARDS (NA, n = 6) and COVID-19 ARDS (CA, n = 7, open circles depicting dexamethasone-treated patients) patients determined by proteomic analysis. Data are mean ± SD. ∗p < 0.05, ∗∗p < 0.01, ∗∗∗p < 0.001, determined by one-way ANOVA and Holm-Sidak’s post hoc-testing. (B) Copy numbers of EIF2AK in the same samples described in (A). Data are mean ± SD. ∗p < 0.05, determined by Kruskal-Wallis and Dunn’s post hoc-testing. (C) Copy numbers of Mx proteins in the same samples described in (A). Data are mean ± SD. ∗p < 0.05, ∗∗p < 0.01, determined by one-way ANOVA and Holm-Sidak’s post hoc-testing. (D) Copy numbers of PSMB proteins in the same samples described in (A). Data are mean ± SD. (E) Volcano plots (left) obtained from proteomic survey of normal density neutrophils (NDN) isolated from the samples described in (A) including a selection of proteins implicated in antigen processing and presentation and interferon signalling (GO:0002376) highlighted in red and with some of them labelled as illustrative examples. Refer to methods section for details. Proteins with a P value <0.05 (horizontal dashed lines), fold change >2 (outer vertical dashed lines) and a copy number >200 in at least one condition after Linear Models for Microarray data analysis were considered as significantly different in the comparisons CA vs HC (top) and NA vs HC (bottom). Table (right) includes a selection of proteins from the samples described in (A) involved in antigen processing and presentation or interferon signalling which significantly change in abundance in CA vs. HC (>2 fold change, p<0.05). Proteins highlighted in red show a COVID-19 specific signature and did not significantly change in non-COVID-19 ARDS alone. Mean copy numbers are shown. (F) Copy numbers of Transporter Associated with Antigen Processing (TAP) proteins in the same samples described in (A). For TAP-1, data are mean ± SD. ∗∗p < 0.01, determined by one-way ANOVA and Holm-Sidak’s post hoc-testing. For TAP-2, data are mean ± SD. ∗p < 0.05, ∗∗p < 0.01, determined by Kruskal-Wallis and Dunn’s post hoc-testing. (G) Copy numbers of Major Histocompatibility Complex molecules in the same samples described in (A). Data are mean ± SD. ∗p < 0.05, ∗∗p < 0.01 determined by one-way ANOVA and Holm-Sidak’s post hoc-testing.

Another important effect of IFN signalling in viral infection is to stimulate antigen presentation of intracellular (i.e. viral) antigens via the proteosome to alert T-cells to the infected cell. Analysis of the antigen presentation and processing pathway showed preserved levels of the immunoproteasome subunits in COVID-19 neutrophils (Figure 7D), but a global increase in the expression of proteins implicated in immune cell development, regulation, antigen processing and presentation (Figure 7E). These included a greater copy number of the Transporter Associated with Antigen Processing (TAP) proteins required for transport into the endoplasmic reticulum for loading onto class I MHC molecules (Figure 7F), and in class I MHC molecules themselves (Figure 7G).

### ARDS neutrophils and type I interferon stimulated healthy neutrophils have altered metabolic profiles with enhanced glutamine utilisation

Type I IFNs have been found to drive metabolic adaptations in plasmacytoid dendritic cells (pDC) with upregulation of fatty acid oxidation and oxidative phosphorylation promoting pDC activation in response to Toll-Like Receptor (TLR) 9 agonists (41). To address whether neutrophils have the capacity to adapt their metabolic programs in response to type I IFN or TLR 7 and 8 activation, blood neutrophils from healthy controls were stimulated in the presence or absence of resiquimod, IFNα and IFNβ and glycolysis was assessed by extracellular flux analysis (Figure 8A). Resiquimod induced a significant uplift in glycolysis and glycolytic capacity, which was partially abrogated by the addition of type I IFNs (Figure 8A). To further characterise the metabolic rewiring in response to type I IFN we undertook LC-MS quantification of individual metabolic intermediaries. In keeping with the real time reduction in extracellular acidification rates, IFN treated neutrophils showed a reduced lactate content (Figure 8B). This was associated with preservation of TCA cycle intermediaires including citrate, alphaketoglutarate, malate and succinate (Figure 8C) and a significant increase in the amino acid glutamate (Figure 8D). Together with (U)-13C5 glutamine tracing into glutamate this would support the ability of neutrophils to substrate switch in response to exposure to type I IFN (Figure 8E). To address whether this metabolic rewiring was observed in blood neutrophils isolated from patients with COVID-19 and non-COVID-19 ARDS we undertook LC-MS analysis of freshly isolated cells. In contrast to type I IFN stimulation of healthy control neutrophils, neutrophils from COVID-19 and non-COVID-19 ARDS patients demonstrated an increase in intracellular levels of free glucose (Figure 8F) while preserving their lactate content (Figure 8G) and TCA cycle intermediaries (Figure 8H) compared to healthy control neutrophils, suggesting these cells have equivalent glycolytic flux and TCA cycle activity. However, in keeping with the capacity of ARDS neutrophils to substrate switch, glutamate levels were elevated in both COVID-19 and non-COVID ARDS neutrophils (Figure 8I).

**Figure 8.**
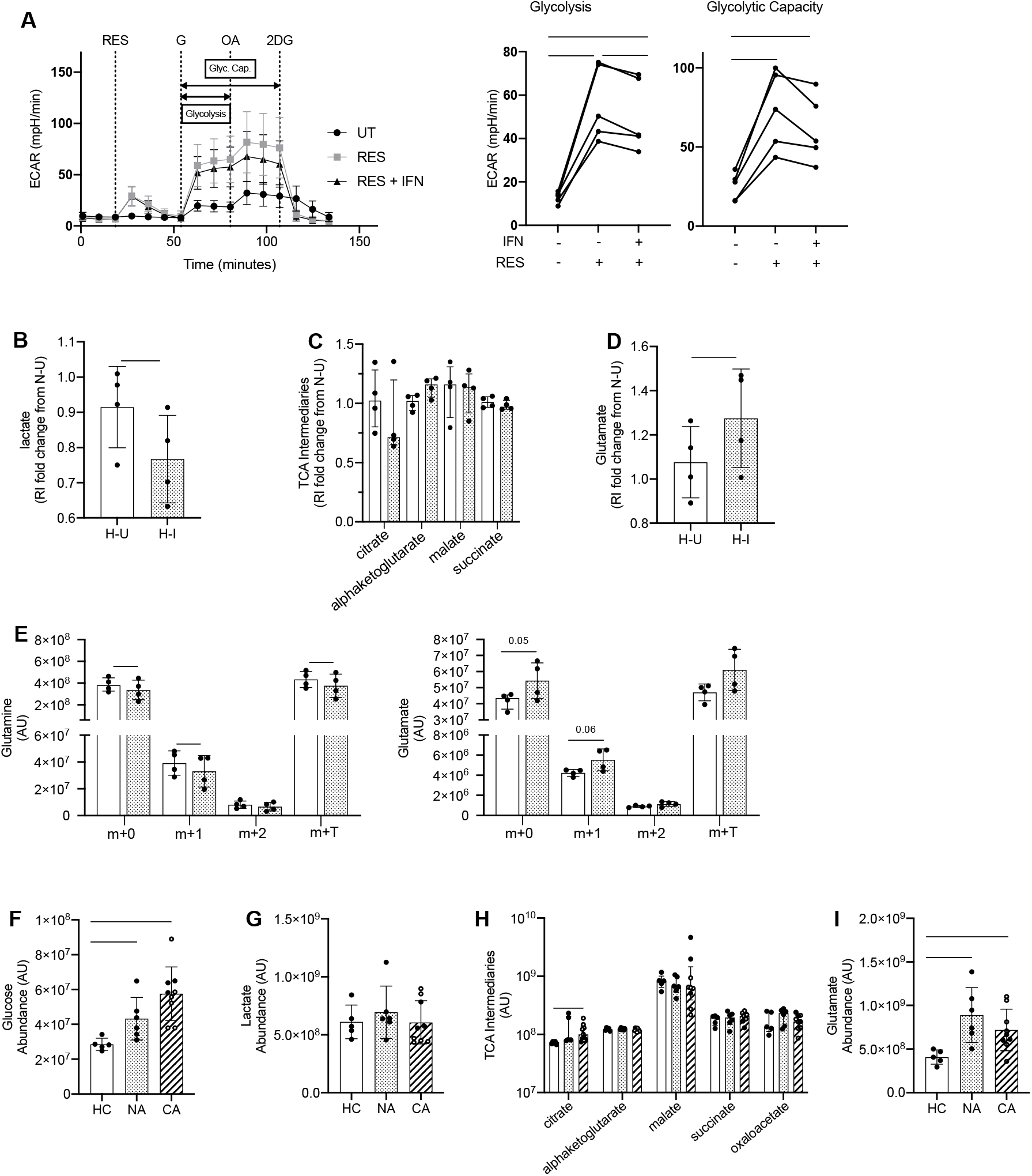
Metabolic rewiring of COVID-19 ARDS neutrophils correspond to changes in neutrophil metabolism in response to type I interferon. (A) Glycolytic behaviour as determined by extracellular flux analysis during the glycolysis stress test in hypoxia (3% O_2_). HC NDN were cultured in hypoxia (1% O_2_) with IFNα/ IFNβ (IFN) or without (UT) for 3 h before sequential injections of resiquimod (RES), glucose (G), oligomycin A (OA) and 2-deoxyglucose (2DG) and extracellular acidification rate (ECAR) determined and used to calculate glycolysis and glycolytic capacity (Glyc. Cap.). Data are mean ± SD (n = 5, individual data points represent mean of at least two technical replicates from individual donors). ∗p < 0.05, ∗∗p < 0.01, determined by repeated measures one ANOVA and Tukey’s post hoc-testing. (B-D) Lactate (B), TCA intermediaries (C) and glutamate (D) abundance in neutrophils cultured in hypoxia for 4 hours with IFNα/ IFNβ (H-I/spotted bars) or without (H-U/open bars) relative to normoxic untreated control cells and determined by p-HILIC LCMS. Data are mean ± SD (n = 4, individual data points represent individual donors). ∗p < 0.05, ∗∗p < 0.01 determined by ratio paired t-test. (E) Glutamine and glutamate isotopomer abundance (Arbitrary Units, AU) as determined by p-HILIC LCMS in neutrophils cultured as in (B-D) in the presence of 2 mM U-13C5 glutamine for 4 h. Individual data points represent individual donors, n = 4. Data are mean ± SD. ∗p<0.05, determined by paired t-test. (F-I) D-glucose (F), lactate (G), TCA intermediary (H) and glutamate (I) abundance in normal density neutrophils (NDN) isolated from healthy controls (HC/open bars, n= 5) and patients with non-COVID-19 ARDS (NA/spotted bars, n = 6) or COVID-19 ARDS (CA/diagonal striped bars, n = 9 with 6 patients receiving dexamethasone treatment indicated by open symbols) was determined by IP LCMS (arbitrary units, AU). Data are mean ± SD (F, G and I) or median ± I.Q.R (H). HC, NA and CA were compared by student’s t-test (F, G and I) or Mann-Whitney (H) where ∗p < 0.05 and ∗∗p < 0.01.

## Discussion

ARDS continues to result in significant mortality despite considerable research endeavour. The emergence of SARS-CoV-2 infection has confounded this, with 10-20% of hospitalised patients progressing to ARDS (42). Urgent understanding of the immunological features specific to COVID-19 ARDS is therefore required. Moreover, the pathophysiological consequences of myeloid dysfunction as determined by scRNA-seq, mass flow cytometry and blood count studies is as yet unclear, as is the mechanism by which dexamethasone improves clinical outcomes in COVID-19 ARDS (14, 40, 43, 44). We therefore employed flow cytometry and mass spectrometry to characterise disease specific protein and metabolite signatures in ARDS neutrophil populations and explored their functional implications. Using this approach, we identify that the expansion of low density and normal density neutrophil populations previously observed in COVID-19 is also observed in non-COVID-19 ARDS. Whilst total cell counts are retained in patients receiving dexamethasone therapy, we report an associated contraction of immature LDN neutrophil populations. It will be important to understand moving forwards whether a key therapeutic effect of dexamethasone is the suppression of acute myelopoiesis in response to infection with SARS-CoV-2.

Proteomic survey also allowed us to highlight key processes, including activation of type I IFN responses that are distinct to COVID-19 ARDS, but more notably, processes not previously detected by scRNA-seq including platelet degranulation and the expression of proteins implicated in immune cell development, regulation, antigen processing and presentation.

Importantly, these protein signatures were observed within mature NDN, suggesting this not to be a consequence of disordered myelopoiesis. It is interesting to note that MCM proteins, that are responsible for the separation of DNA and as such are conventionally associated with DNA replication, were enriched in both COVID-19 ARDS and non-COVID-19 ARDS. In the context of neutrophils, a terminally differentiated cell, this may hint to processes involved in the unravelling of DNA for NET formation rather than cell cycle control and serves as an interesting concept for further exploration.

A striking clinical divergence between COVID-19 and non-COVID-19 ARDS is the prominence of micro and macrovascular thrombosis in COVID-19 ARDS. Here, we report proteomic signatures indicative of platelet degranulation and clotting cascade activation. These observations together with evidence of neutrophil platelet binding, extend the previously reported contribution of neutrophils to the pathogenesis of immune clot formation through the release of NETs to one of TLR mediated neutrophil activation and the formation of neutrophil platelet aggregates. It is interesting to note that it is the neutrophils within the low density layer we observe by confocal microscopy to bind to platelets and to be associated with the upregulating the Mac-1 platelet binding complex in COVID-19 ARDS. Further work will be required to understand whether LDN also demonstrate a propensity for NETs formation, and whether these aggregates, previously reported in the lung tissue at post-mortem, impair neutrophil transmigration and directly contribute to vascular damage and to the formation of microthrombi (17-19). It will also be important to dissect whether the uplift in expression of proteins associated with fibrin clot formation in COVID-19 ARDS is consequent upon intrinsic neutrophil expression of these proteins, neutrophil processing of platelet proteins or reflective of adherent platelets contributing to the protein signatures of the circulating neutrophil populations.

The importance of neutrophil activation of type I IFN signalling pathways in COVID-19 ARDS also requires further consideration given the disconnect between tissue injury and viral detection (45). The ability of neutrophils to cross-present exogenous antigens to CD8+ T cells has previously been reported and is highly relevant for T cell priming in vivo (46). This may be particularly relevant in a disease where early CD4+ and CD8+ T cell responses against SARS-CoV-2 are thought to be protective (47, 48), but late responses are associated with damaging inflammation (48-51). Whilst activation of anti-viral responses including class I MHC antigen presentation would therefore appear beneficial with respect to viral control, if this is associated with a hyper-inflammatory neutrophil phenotype and delayed T cell activation, the net consequence could be persistent tissue injury. In this regard, we would predict that inappropriate degranulation of neutrophils in the circulation would be highly damaging and cause wide-spread inflammation within the microvasculature where neutrophils are known to be sequestered. Our evidence of expanded neutrophil numbers together with increased neutrophil activation and degranulation confirmed by the detection of plasma neutrophil granule proteins in patients with COVID-19 ARDS and TLR-agonist in vitro assays would support this concept of a hyper-inflammatory, damaging circulating innate response. These pro-thrombotic hyperinflammatory neutrophil protein signatures would appear to be retained in patients with COVID-19 ARDS receiving dexamethasone therapy. This is perhaps not surprising given the paucity of evidence for the impact of corticosteroids on neutrophilic inflammation, highlighting the need for additional therapeutic strategies.

Finally, the mechanism by which type I IFN regulates neutrophil behaviour remains to be fully elucidated. In pDC, TLR 9 mediated activation is dependent upon autocrine production of type I IFNs and an increase in oxidative metabolism (41). Neutrophils are unique in their reliance on non-oxidative metabolism for ATP production, even when oxygen is freely available. It is therefore of interest that in response to IFNα and IFNβ, neutrophils rewire their metabolic programme by reducing their glycolytic potential. Together with an increase in detectable levels of glutamate in neutrophils isolated from patients with COVID-19 ARDS, and existing evidence that neutrophils can undergo gluconeogenesis (52), this raises the interesting possibility that ARDS neutrophils re-wire their metabolic processes to facilitate ongoing inflammatory responses which may be detrimental to the host. Future work will be required to better understand whether these metabolic changes potentiate anti-viral and pro-inflammatory innate immune responses following viral challenge. Of interest small molecules already exist for targeting glutamine utilisation and have been trialled in the cancer setting, raising the possibility of metabolic drug repurposing for the treatment of COVID-19 (53).

In summary, using mass spectrometry we describe pathophysiological protein and metabolic neutrophil signatures that are common to ARDS and those distinct to COVID-19 ARDS. We identify a type I IFN response in COVID-19 ARDS neutrophils which is associated with metabolic rewiring, neutrophil degranulation and the formation of neutrophil platelet aggregates in the blood which persist irrespective of dexamethasone treatment. A clear limitation of this study is the relatively small number of patients recruited. This is balanced against the detailed analysis we have been able to perform in these patient groups and the move towards functional dissection of neutrophil responses not previously captured in transcriptional data sets. Further understanding of the mechanisms which regulate aberrant neutrophil responses will likely be important in developing strategies to target the innate responses following infection with SARS-CoV-2 to enable an effective therapeutic arsenal for COVID-19 ARDS.

## Data Availability

Raw mass spectrometry data files and Spectronaut analysis files will be available to download from the ProteomeXchange data repository (http://proteomecentral.proteomexchange.org/cgi/GetDataset) at the time of publication.

## Author Contributions

L.R, M.A.S.G, T.M, A.J.M.H, E.R.W, S.A, P.S, P.C, A.S.M, A.Z, D.H, S.K.C, J.S, S.J, R.G, A.P, S.M, I.H, M.H.F, A.B, S.P, A.L, G.R.B, B.G, W.V, C.T.C performed the research. L.R, M.A.S.G, T.M, A.J.M.H, M.K.W, D.G, D.A.C, S.R.W interpreted the data. L.R, M.A.S.G, T.M, A.J.M.H, M.H.F, K.D, N.H, D.D, M.K.W, D.G, D.A.C, S.R.W designed the research. L.R, M.A.S.G, T.M, A.J.M.H, E.R.W, A.K., M.K.W, D.G, D.A.C, S.R.W provided expertise and feedback. L.R, M.A.S.G, T.M, A.J.M.H, E.R.W, D.A.C, S.R.W wrote the manuscript.

## Acknowledgements

We thank the CIR blood resource (AMREC no. 15-HV-013) for the recruitment of blood from healthy donors and the clinical support teams, patients and their families who have contributed to this study. Many thanks to the QMRI Flow Cytometry & Cell Sorting Facility, Edinburgh University (Will Ramsay and Mari Pattison) and CALM Facility, Edinburgh University (Rolly Wiegand and Kseniya Korobchevskaya) for their expertise and assistance.

## References

1. Huang C, Wang Y, Li X, Ren L, Zhao J, Hu Y, Zhang L, Fan G, Xu J, Gu X, Cheng Z, Yu T, Xia J, Wei Y, Wu W, Xie X, Yin W, Li H, Liu M, Xiao Y, Gao H, Guo L, Xie J, Wang G, Jiang R, Gao Z, Jin Q, Wang J, Cao B. Clinical features of patients infected with 2019 novel coronavirus in Wuhan, China. Lancet 2020; 395: 497–506.

2. Acute Respiratory Distress Syndrome Network, Brower RG, Matthay MA, Morris A, Schoenfeld D, Thompson BT, Wheeler A. Ventilation with lower tidal volumes as compared with traditional tidal volumes for acute lung injury and the acute respiratory distress syndrome. N Engl J Med 2000; 342: 1301–1308.

3. Dreyfuss D, Saumon G. Role of tidal volume, FRC, and end-inspiratory volume in the development of pulmonary edema following mechanical ventilation. Am Rev Respir Dis 1993; 148: 1194–1203.

4. Bachofen M, Weibel ER. Alterations of the gas exchange apparatus in adult respiratory insufficiency associated with septicemia. Am Rev Respir Dis 1977; 116: 589–615.

5. Flick MR, Perel A, Staub NC. Leukocytes are required for increased lung microvascular permeability after microembolization in sheep. Circ Res 1981; 48: 344–351.

6. Tian S, Xiong Y, Liu H, Niu L, Guo J, Liao M, Xiao SY. Pathological study of the 2019 novel coronavirus disease (COVID-19) through postmortem core biopsies. Mod Pathol 2020; 33: 1007–1014.

7. Sharp C, Millar AB, Medford AR. Advances in understanding of the pathogenesis of acute respiratory distress syndrome. Respiration 2015; 89: 420–434.

8. Eltzschig HK, Carmeliet P. Hypoxia and inflammation. N Engl J Med 2011; 364: 656–665.

9. Walmsley SR, Print C, Farahi N, Peyssonnaux C, Johnson RS, Cramer T, Sobolewski A, Condliffe AM, Cowburn AS, Johnson N, Chilvers ER. Hypoxia-induced neutrophil survival is mediated by HIF-1alpha-dependent NF-kappaB activity. J Exp Med 2005; 201: 105–115.

10. Tobin MJ. Basing Respiratory Management of COVID-19 on Physiological Principles. Am J Respir Crit Care Med 2020; 201: 1319–1320.

11. Liu J, Liu Y, Xiang P, Pu L, Xiong H, Li C, Zhang M, Tan J, Xu Y, Song R, Song M, Wang L, Zhang W, Han B, Yang L, Wang X, Zhou G, Zhang T, Li B, Wang Y, Chen Z, Wang X. Neutrophil-to-lymphocyte ratio predicts critical illness patients with 2019 coronavirus disease in the early stage. J Transl Med 2020; 18: 206.

12. Zhao Q, Meng M, Kumar R, Wu Y, Huang J, Deng Y, Weng Z, Yang L. Lymphopenia is associated with severe coronavirus disease 2019 (COVID-19) infections: A systemic review and meta-analysis. Int J Infect Dis 2020; 96: 131–135.

13. Dorward DA, Russell CD, Um IH, Elshani M, Armstrong SD, Penrice-Randal R, Millar T, Lerpiniere CEB, Tagliavini G, Hartley CS, Randle NP, Gachanja NN, Potey PMD, Dong X, Anderson AM, Campbell VL, Duguid AJ, Al Qsous W, BouHaidar R, Baillie JK, Dhaliwal K, Wallace WA, Bellamy COC, Prost S, Smith C, Hiscox JA, Harrison DJ, Lucas CD. Tissue-Specific Immunopathology in Fatal COVID-19. Am J Respir Crit Care Med 2021; 203: 192–201.

14. Schulte-Schrepping J, Reusch N, Paclik D, Bassler K, Schlickeiser S, Zhang B, Kramer B, Krammer T, Brumhard S, Bonaguro L, De Domenico E, Wendisch D, Grasshoff M, Kapellos TS, Beckstette M, Pecht T, Saglam A, Dietrich O, Mei HE, Schulz AR, Conrad C, Kunkel D, Vafadarnejad E, Xu CJ, Horne A, Herbert M, Drews A, Thibeault C, Pfeiffer M, Hippenstiel S, Hocke A, Muller-Redetzky H, Heim KM, Machleidt F, Uhrig A, Bosquillon de Jarcy L, Jurgens L, Stegemann M, Glosenkamp CR, Volk HD, Goffinet C, Landthaler M, Wyler E, Georg P, Schneider M, Dang-Heine C, Neuwinger N, Kappert K, Tauber R, Corman V, Raabe J, Kaiser KM, Vinh MT, Rieke G, Meisel C, Ulas T, Becker M, Geffers R, Witzenrath M, Drosten C, Suttorp N, von Kalle C, Kurth F, Handler K, Schultze JL, Aschenbrenner AC, Li Y, Nattermann J, Sawitzki B, Saliba AE, Sander LE, Deutsche C-OI. Severe COVID-19 Is Marked by a Dysregulated Myeloid Cell Compartment. Cell 2020.

15. Yang P, Li Y, Xie Y, Liu Y. Different Faces for Different Places: Heterogeneity of Neutrophil Phenotype and Function. J Immunol Res 2019; 2019: 8016254.

16. Group RC, Horby P, Lim WS, Emberson JR, Mafham M, Bell JL, Linsell L, Staplin N, Brightling C, Ustianowski A, Elmahi E, Prudon B, Green C, Felton T, Chadwick D, Rege K, Fegan C, Chappell LC, Faust SN, Jaki T, Jeffery K, Montgomery A, Rowan K, Juszczak E, Baillie JK, Haynes R, Landray MJ. Dexamethasone in Hospitalized Patients with Covid-19 - Preliminary Report. N Engl J Med 2020.

17. Nicolai L, Leunig A, Brambs S, Kaiser R, Weinberger T, Weigand M, Muenchhoff M, Hellmuth JC, Ledderose S, Schulz H, Scherer C, Rudelius M, Zoller M, Hochter D, Keppler O, Teupser D, Zwissler B, Bergwelt-Baildon M, Kaab S, Massberg S, Pekayvaz K, Stark K. Immunothrombotic Dysregulation in COVID-19 Pneumonia is Associated with Respiratory Failure and Coagulopathy. Circulation 2020.

18. Radermecker C, Detrembleur N, Guiot J, Cavalier E, Henket M, d’Emal C, Vanwinge C, Cataldo D, Oury C, Delvenne P, Marichal T. Neutrophil extracellular traps infiltrate the lung airway, interstitial, and vascular compartments in severe COVID-19. Journal of Experimental Medicine 2020; 217.

19. Veras FP, Pontelli MC, Silva CM, Toller-Kawahisa JE, de Lima M, Nascimento DC, Schneider AH, Caetité D, Tavares LA, Paiva IM, Rosales R, Colón D, Martins R, Castro IA, Almeida GM, Lopes MIF, Benatti MN, Bonjorno LP, Giannini MC, Luppino-Assad R, Almeida SL, Vilar F, Santana R, Bollela VR, Auxiliadora-Martins M, Borges M, Miranda CH, Pazin-Filho A, da Silva LLP, Cunha L, Zamboni DS, Dal-Pizzol F, Leiria LO, Siyuan L, Batah S, Fabro A, Mauad T, Dolhnikoff M, Duarte-Neto A, Saldiva P, Cunha TM, Alves-Filho JC, Arruda E, Louzada-Junior P, Oliveira RD, Cunha FQ. SARS-CoV-2–triggered neutrophil extracellular traps mediate COVID-19 pathology. Journal of Experimental Medicine 2020; 217.

20. ARDS Defence Task Force, Ranieri VM, Rubenfeld GD, Thompson BT, Ferguson ND, Caldwell E, Fan E, Camporota L, Slutsky AS. Acute respiratory distress syndrome: the Berlin Definition. JAMA 2012; 307: 2526–2533.

21. Knaus WA, Draper EA, Wagner DP, Zimmerman JE. APACHE II: a severity of disease classification system. Crit Care Med 1985; 13: 818–829.

22. Groll DL, To T, Bombardier C, Wright JG. The development of a comorbidity index with physical function as the outcome. J Clin Epidemiol 2005; 58: 595–602.

23. Haslett C, Guthrie LA, Kopaniak MM, Johnston RB, Jr., Henson PM. Modulation of multiple neutrophil functions by preparative methods or trace concentrations of bacterial lipopolysaccharide. Am J Pathol 1985; 119: 101–110.

24. Walmsley SR, Chilvers ER, Thompson AA, Vaughan K, Marriott HM, Parker LC, Shaw G, Parmar S, Schneider M, Sabroe I, Dockrell DH, Milo M, Taylor CT, Johnson RS, Pugh CW, Ratcliffe PJ, Maxwell PH, Carmeliet P, Whyte MK. Prolyl hydroxylase 3 (PHD3) is essential for hypoxic regulation of neutrophilic inflammation in humans and mice. J Clin Invest 2011; 121: 1053–1063.

25. HaileMariam M, Eguez RV, Singh H, Bekele S, Ameni G, Pieper R, Yu Y. S-Trap, an Ultrafast Sample-Preparation Approach for Shotgun Proteomics. J Proteome Res 2018; 17: 2917–2924.

26. Muntel J, Gandhi T, Verbeke L, Bernhardt OM, Treiber T, Bruderer R, Reiter L. Surpassing 10 000 identified and quantified proteins in a single run by optimizing current LC-MS instrumentation and data analysis strategy. Mol Omics 2019; 15: 348– 360.

27. Bruderer R, Bernhardt OM, Gandhi T, Miladinovic SM, Cheng LY, Messner S, Ehrenberger T, Zanotelli V, Butscheid Y, Escher C, Vitek O, Rinner O, Reiter L. Extending the limits of quantitative proteome profiling with data-independent acquisition and application to acetaminophen-treated three-dimensional liver microtissues. Mol Cell Proteomics 2015; 14: 1400–1410.

28. Ritchie ME, Phipson B, Wu D, Hu Y, Law CW, Shi W, Smyth GK. limma powers differential expression analyses for RNA-sequencing and microarray studies. Nucleic Acids Res 2015; 43: e47.

29. Wisniewski JR, Hein MY, Cox J, Mann M. A “proteomic ruler” for protein copy number and concentration estimation without spike-in standards. Mol Cell Proteomics 2014; 13: 3497–3506.

30. Schindelin J, Arganda-Carreras I, Frise E, Kaynig V, Longair M, Pietzsch T, Preibisch S, Rueden C, Saalfeld S, Schmid B, Tinevez JY, White DJ, Hartenstein V, Eliceiri K, Tomancak P, Cardona A. Fiji: an open-source platform for biological-image analysis. Nat Methods 2012; 9: 676–682.

31. WHO. Clinical management of COVID-19: interim guidance. WHO Global; 2020.

32. Silvestre-Roig C, Fridlender ZG, Glogauer M, Scapini P. Neutrophil Diversity in Health and Disease. Trends Immunol 2019; 40: 565–583.

33. Diebold SS, Kaisho T, Hemmi H, Akira S, Reis e Sousa C. Innate antiviral responses by means of TLR7-mediated recognition of single-stranded RNA. Science 2004; 303: 1529–1531.

34. Moreno-Eutimio MA, Lopez-Macias C, Pastelin-Palacios R. Bioinformatic analysis and identification of single-stranded RNA sequences recognized by TLR7/8 in the SARS-CoV-2, SARS-CoV, and MERS-CoV genomes. Microbes Infect 2020; 22: 226–229.

35. Hattermann K, Picard S, Borgeat M, Leclerc P, Pouliot M, Borgeat P. The Toll-like receptor 7/8-ligand resiquimod (R-848) primes human neutrophils for leukotriene B4, prostaglandin E2 and platelet-activating factor biosynthesis. FASEB J 2007; 21: 1575-1585.

36. Grommes J, Soehnlein O. Contribution of neutrophils to acute lung injury. Mol Med 2011; 17: 293–307.

37. Vogt KL, Summers C, Chilvers ER, Condliffe AM. Priming and de-priming of neutrophil responses in vitro and in vivo. Eur J Clin Invest 2018; 48 Suppl 2: e12967.

38. Hung IF, Lung KC, Tso EY, Liu R, Chung TW, Chu MY, Ng YY, Lo J, Chan J, Tam AR, Shum HP, Chan V, Wu AK, Sin KM, Leung WS, Law WL, Lung DC, Sin S, Yeung P, Yip CC, Zhang RR, Fung AY, Yan EY, Leung KH, Ip JD, Chu AW, Chan WM, Ng AC, Lee R, Fung K, Yeung A, Wu TC, Chan JW, Yan WW, Chan WM, Chan JF, Lie AK, Tsang OT, Cheng VC, Que TL, Lau CS, Chan KH, To KK, Yuen KY. Triple combination of interferon beta-1b, lopinavir-ritonavir, and ribavirin in the treatment of patients admitted to hospital with COVID-19: an open-label, randomised, phase 2 trial. Lancet 2020; 395: 1695–1704.

39. Lucas C, Wong P, Klein J, Castro TBR, Silva J, Sundaram M, Ellingson MK, Mao T, Oh JE, Israelow B, Takahashi T, Tokuyama M, Lu P, Venkataraman A, Park A, Mohanty S, Wang H, Wyllie AL, Vogels CBF, Earnest R, Lapidus S, Ott IM, Moore AJ, Muenker MC, Fournier JB, Campbell M, Odio CD, Casanovas-Massana A, Yale IT, Herbst R, Shaw AC, Medzhitov R, Schulz WL, Grubaugh ND, Dela Cruz C, Farhadian S, Ko AI, Omer SB, Iwasaki A. Longitudinal analyses reveal immunological misfiring in severe COVID-19. Nature 2020; 584: 463–469.

40. Wu D, Sanin DE, Everts B, Chen Q, Qiu J, Buck MD, Patterson A, Smith AM, Chang CH, Liu Z, Artyomov MN, Pearce EL, Cella M, Pearce EJ. Type 1 Interferons Induce Changes in Core Metabolism that Are Critical for Immune Function. Immunity 2016; 44: 1325–1336.

41. Quah P, Li A, Phua J. Mortality rates of patients with COVID-19 in the intensive care unit: a systematic review of the emerging literature. Crit Care 2020; 24: 285.

42. Mann ER, Menon M, Knight SB, Konkel JE, Jagger C, Shaw TN, Krishnan S, Rattray M, Ustianowski A, Bakerly ND, Dark P, Lord G, Simpson A, Felton T, Ho LP, TRC NR, Feldmann M, Circo, Grainger JR, Hussell T. Longitudinal immune profiling reveals key myeloid signatures associated with COVID-19. Sci Immunol 2020; 5.

43. Silvin A, Chapuis N, Dunsmore G, Goubet AG, Dubuisson A, Derosa L, Almire C, Henon C, Kosmider O, Droin N, Rameau P, Catelain C, Alfaro A, Dussiau C, Friedrich C, Sourdeau E, Marin N, Szwebel TA, Cantin D, Mouthon L, Borderie D, Deloger M, Bredel D, Mouraud S, Drubay D, Andrieu M, Lhonneur AS, Saada V, Stoclin A, Willekens C, Pommeret F, Griscelli F, Ng LG, Zhang Z, Bost P, Amit I, Barlesi F, Marabelle A, Pene F, Gachot B, Andre F, Zitvogel L, Ginhoux F, Fontenay M, Solary E. Elevated Calprotectin and Abnormal Myeloid Cell Subsets Discriminate Severe from Mild COVID-19. Cell 2020.

44. Dorward DA, Russell CD, Um IH, Elshani M, Armstrong SD, Penrice-Randal R, Millar T, Lerpiniere CE, Tagliavini G, Hartley CS, Randall NP, Gachanja NN, Potey PM, Anderson AM, Campbell VL, Duguid AJ, Al Qsous W, BouHaidar R, Baillie JK, Dhaliwal K, Wallace WA, Bellamy CO, Prost S, Smith C, Hiscox JA, Harrison DJ, Lucas CD,. Tissue-specific tolerance in fatal Covid-19. medRxiv 2020.

45. Pufnock JS, Cigal M, Rolczynski LS, Andersen-Nissen E, Wolfl M, McElrath MJ, Greenberg PD. Priming CD8+ T cells with dendritic cells matured using TLR4 and TLR7/8 ligands together enhances generation of CD8+ T cells retaining CD28. Blood 2011; 117: 6542–6551.

46. Blanco-Melo D, Nilsson-Payant BE, Liu WC, Uhl S, Hoagland D, Moller R, Jordan TX, Oishi K, Panis M, Sachs D, Wang TT, Schwartz RE, Lim JK, Albrecht RA, tenOever BR. Imbalanced Host Response to SARS-CoV-2 Drives Development of COVID-19. Cell 2020; 181: 1036–1045 e1039.

47. Grifoni A, Weiskopf D, Ramirez SI, Mateus J, Dan JM, Moderbacher CR, Rawlings SA, Sutherland A, Premkumar L, Jadi RS, Marrama D, de Silva AM, Frazier A, Carlin AF, Greenbaum JA, Peters B, Krammer F, Smith DM, Crotty S, Sette A. Targets of T Cell Responses to SARS-CoV-2 Coronavirus in Humans with COVID-19 Disease and Unexposed Individuals. Cell 2020; 181: 1489–1501 e1415.

48. Chen G, Wu D, Guo W, Cao Y, Huang D, Wang H, Wang T, Zhang X, Chen H, Yu H, Zhang X, Zhang M, Wu S, Song J, Chen T, Han M, Li S, Luo X, Zhao J, Ning Q. Clinical and immunological features of severe and moderate coronavirus disease 2019. J Clin Invest 2020; 130: 2620–2629.

49. Li CK, Wu H, Yan H, Ma S, Wang L, Zhang M, Tang X, Temperton NJ, Weiss RA, Brenchley JM, Douek DC, Mongkolsapaya J, Tran BH, Lin CL, Screaton GR, Hou JL, McMichael AJ, Xu XN. T cell responses to whole SARS coronavirus in humans. J Immunol 2008; 181: 5490–5500.

50. Liu L, Wei Q, Lin Q, Fang J, Wang H, Kwok H, Tang H, Nishiura K, Peng J, Tan Z, Wu T, Cheung KW, Chan KH, Alvarez X, Qin C, Lackner A, Perlman S, Yuen KY, Chen Z. Anti-spike IgG causes severe acute lung injury by skewing macrophage responses during acute SARS-CoV infection. JCI Insight 2019; 4.

51. Sadiku P, Willson JA, Ryan EM, Sammut D, Coelho P, Watts ER, Grecian R, Young JM, Bewley M, Arienti S, Mirchandani AS, Sanchez Garcia MA, Morrison T, Zhang A, Reyes L, Griessler T, Jheeta P, Paterson GG, Graham CJ, Thomson JP, Baillie K, Thompson AAR, Morgan JM, Acosta-Sanchez A, Darde VM, Duran J, Guinovart JJ, Rodriguez-Blanco G, Von Kriegsheim A, Meehan RR, Mazzone M, Dockrell DH, Ghesquiere B, Carmeliet P, Whyte MKB, Walmsley SR. Neutrophils Fuel Effective Immune Responses through Gluconeogenesis and Glycogenesis. Cell Metab 2020.

52. McBrayer SK, Mayers JR, DiNatale GJ, Shi DD, Khanal J, Chakraborty AA, Sarosiek KA, Briggs KJ, Robbins AK, Sewastianik T, Shareef SJ, Olenchock BA, Parker SJ, Tateishi K, Spinelli JB, Islam M, Haigis MC, Looper RE, Ligon KL, Bernstein BE, Carrasco RD, Cahill DP, Asara JM, Metallo CM, Yennawar NH, Vander Heiden MG, Kaelin WG, Jr. Transaminase Inhibition by 2-Hydroxyglutarate Impairs Glutamate Biosynthesis and Redox Homeostasis in Glioma. Cell 2018; 175: 101–116 e125.

